# Clinical long-read genome sequencing for rare disease diagnostics

**DOI:** 10.64898/2026.01.13.26343759

**Authors:** Tessa J.J. de Bitter, Bart van der Sanden, Lydia Sagath, Wolfram Höps, Peer Arts, Michelle de Groot, Marjan M. Weiss, Ronny Derks, Amber den Ouden, Simone van den Heuvel, Raoul G.J. Timmermans, Timon van Leeuwen, Jordi Corominas Galbany, Jos Smits, Lot Snijders Blok, Tom Hofste, Marloes Steehouwer, Nick Zomer, Quentin Sabbagh, Erik-Jan Kamsteeg, Dorien Lugtenberg, Ermanno A. Bosgoed, Richard J. Rodenburg, Su Ming Sun, Arjen R. Mensenkamp, Marjolijn J.L. Ligtenberg, Nicole de Leeuw, Debby M.E.I. Hellebrekers, Alexander P.A. Stegmann, Aimée D.C. Paulussen, Marinus J. Blok, Wendy A.G. van Zelst-Stams, Arthur van den Wijngaard, Helger G. Yntema, Christian Gilissen, Alexander Hoischen, Lisenka E.L.M. Vissers

## Abstract

**Background:** Diagnostic evaluation of rare genetic disorders continues to rely on multiple test modalities, despite the increasing use of short-read exome or genome sequencing as first-tier tests. Long-read genome sequencing (lrGS) has the potential to consolidate current standard-of-care (SoC) diagnostics into a single assay, but its accuracy and clinical utility in routine practice have not been established at large scale.

**Methods:** We studied 1000 clinical samples, including 832 index cases, representative of one year of germline diagnostic testing at two tertiary centers. Samples underwent lrGS at approximately 30× coverage, followed by variant detection and interpretation. Diagnostic outcomes were compared with those obtained through SoC testing. Using these results, we modeled the effect of implementing lrGS as a first-tier test in a cohort of 15,150 index cases who underwent diagnostic testing during a single year.

**Results:** The overall concordance between SoC and lrGS testing was 96.4%. lrGS identified clinically relevant findings that improved or refined genetic diagnoses in 3.4% of cases, largely through phasing for recessive disorders and the detection of novel disease-causing variants. In 0.2% of cases, SoC testing identified variants not detected by lrGS. Modeling of a generic lrGS-first diagnostic strategy demonstrated an estimated absolute increase in diagnostic yield of 2.5% across the full annual cohort.

**Conclusions:** LrGS demonstrated high concordance with current diagnostic approaches and provided incremental diagnostic benefits. These findings support lrGS as a feasible and effective first-tier test for rare disease diagnostics.

## Introduction

Of >7,000 rare diseases, 70–80% are estimated to have a genetic origin^1^. Establishing a molecular diagnosis provides substantial clinical benefits for patients and families, including targeted or precautionary treatment, prognostic insight, and informed genetic counselling. This information can guide clinical decision-making, family planning, and cascade testing of at-risk relatives^2,3^. The introduction of next-generation sequencing has transformed genetic diagnostics for rare diseases. Since 2012, exome sequencing (ES) has been widely adopted as a first-tier test, as its genotype-driven approach helps address the clinical and genetic heterogeneity of rare diseases^4,5^. More recently, short-read genome sequencing (srGS) has further improved the detection of single nucleotide variants and small insertions or deletions (SNV/InDels, <50bp), copy-number variants (CNVs) including structural variants (SVs) and repeat expansions, including comprehensive detection of non-coding variants^6,7^.

While srGS is currently the most complete first-tier test for most rare diseases, its use has not led to dramatically increased diagnostic yields compared with ES, still leaving a majority of patients without a genetic diagnosis^6,8,9^. In part, this is expected to be due to intrinsic limitations of short-read technologies, as they cannot robustly identify all classes of genome variation, requiring complementary technologies for routine detection of all clinically relevant genetic variation for rare diseases^7^. These additional tests increase turn-around times but also require many different technologies. A single generic genetic test could increase efficiency and decrease time-to-diagnosis in diagnostic laboratories.

Long-read genome sequencing (lrGS) technologies, including nanopore sequencing (Oxford Nanopore Technology) and high fidelity (HiFi) sequencing (PacBio), generate sequence reads of tens of kilo bases (kb) in size. These longer reads enable improved SV detection but also haplotype generation for variant phasing, distinguishing clinically relevant genes from their pseudogenes, and determining the length and sequence context of repeat expansions^10,11^. Additionally, lrGS provides direct readouts of 5-methylcytosine (5mC) DNA methylation (DNAm), which can be relevant for the diagnosis of imprinting disorders such as Prader-Willi (MIM 176270) and Angelman syndromes (MIM 105830). It can also detect skewed X-inactivation and help to functionally evaluate germline variants using DNAm profiles^12,13^. Recent improvements in sequencing accuracy have made lrGS comparable to short-read approaches for detecting SNVs/InDels^14,15^ with PacBio HiFi demonstrating superior performance in regions challenging for srGS^16^.

To systematically evaluate lrGS as a comprehensive first-tier diagnostic test, we analyzed a cohort of 1,000 clinical samples representative of annual postnatal germline testing at our medical centers. For each sample, lrGS was performed alongside standard-of-care (SoC) testing. We compared clinical utility and assessed the feasibility and potential impact of implementing HiFi-based lrGS as a generic first-tier diagnostic approach.

## Methods

### Cohort

A set of 1,000 samples was selected from index cases referred for postnatal germline diagnostic testing the department of Human Genetics of the Radboudumc and the department of Clinical Genetics of Maastricht UMC+ in 2022 (**Figure 1A**, **Table 1, Table S1**). The cohort was representative of the centers’ combined annual germline testing with respect to referral volume, diagnostic yield, and use of standard-of-care workflows, each targeting specific variant types or loci (**Table S1**)^7^. Cohort details are provided in **Supplementary Appendix 1**.

**Figure 1.**
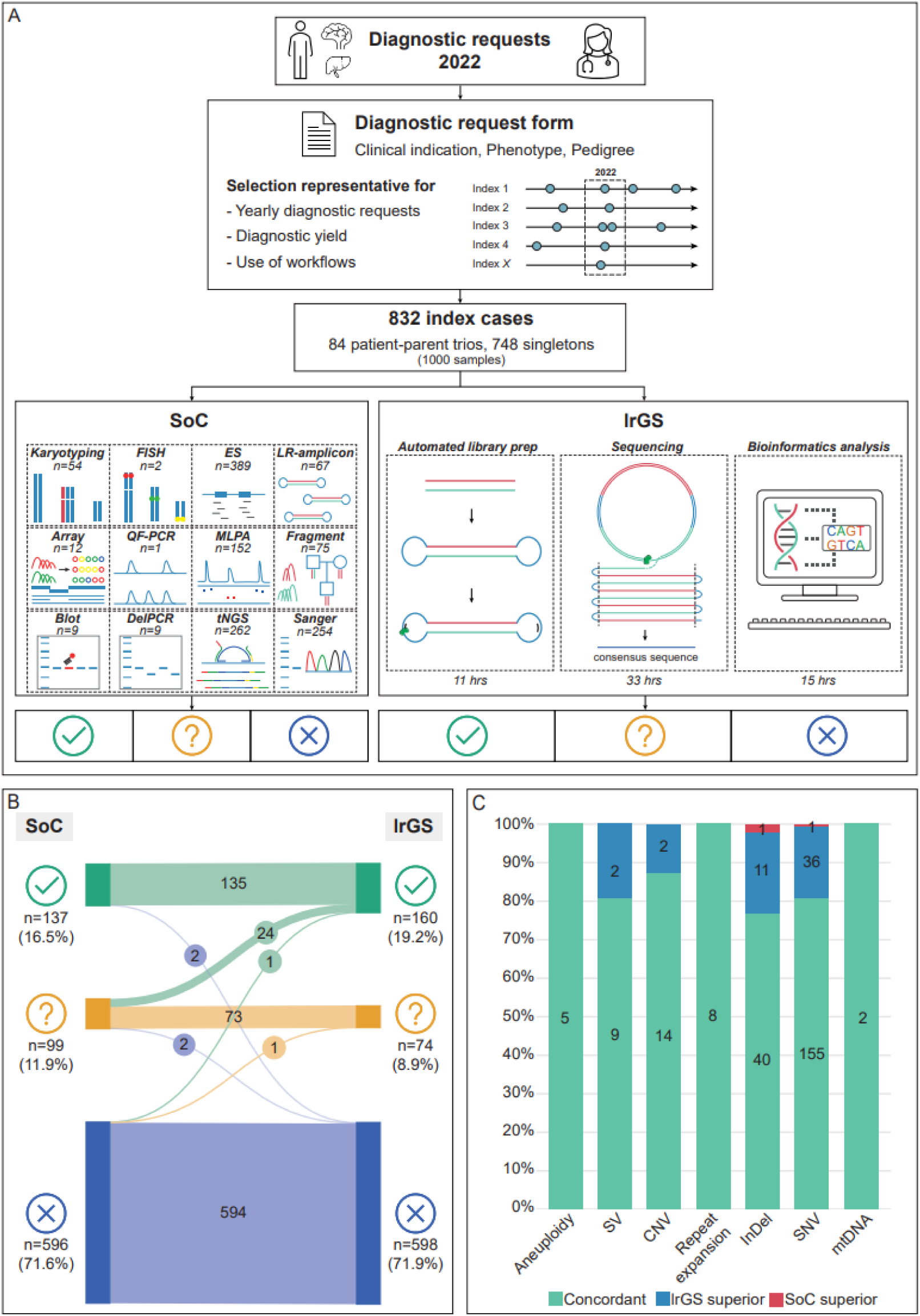
Diagnostic yield comparison to SoC. **A)** Study design and workflow. Details of lrGS bioinformatics pipeline in **Figure S1. B)** Comparison of diagnostic outcomes for SoC and lrGS for 832 index cases at moment of inclusion (**Table S1**). **C)** Concordance for detection of the variant types resulting in conclusive and possible diagnoses in SoC and lrGS at moment of inclusion (**Table S3**). SoC: standard-of-care; lrGS: long-read genome sequencing; FISH: fluorescence in situ hybridisation; LR-amplicon: long-read amplicon; QF-PCR: Quantitative fluorescence PCR; MLPA: multiplex ligation-dependent probe amplification; DelPCR: deletion PCR; tNGS: targeted NGS; SV: structural variant; CNV: copy-number variant; InDel: insertion/deletion; SNV; single-nucleotide variant; mtDNA: mitochondrial DNA.

**Table 1.** cohort demographics.

| **Characteristic** | **Index cases with Sequencing (n=832)** |
| --- | --- |
| **Sex - n (%)** |  |
| Male | 402 (48.3) |
| Female | 430 (51.7) |
| **Median age at included referral - yr (range)** | **37 (0–91)** |
| **Clinical indication - n (%)** |  |
| Metabolic & Mitochondrial disorders | 77 (9.3) |
| Disorders in sex development, Endocrine, Fertility and Pregnancy | 73 (8.8) |
| Thoracic, abdominal and vascular organ systems | 77 (9.3) |
| Hearing and/or Vision impairment | 61 (7.3) |
| Hematological disorders | 29 (3.5) |
| Immunological disorders | 39 (4.7) |
| Neurodevelopmental disorders/multiple congenital anomalies | 65 (7.8) |
| Neurological disorders | 118 (14.2) |
| Neuromuscular disorders | 28 (3.4) |
| Cancer germline predisposition | 140 (16.7) |
| Skeletal diseases | 31 (3.7) |
| Skin disorders | 32 (3.8) |
| Other, complex conditions | 62 (7.5) |
| **Type of analysis - n (%)** |  |
| Singleton | 748 (89.9) |
| Trio | 84 (10.1) |
| **Standard of Care at moment of inclusion** |  |
| Average number of genetic requests – n (range) | 1.21 (1–5) |
| Targeted approach – n (%)<br>(FISH, LR-amplicon, QF-PCR, MLPA, Fragment analysis, Blot, DelPCR, tNGS, Sanger) | 424 (51.0) |
| Genome-wide approach – n (%)<br>(Karyotyping, ES, Array) | 359 (43.1) |
| Combination of targeted and genome-wide approach – n (%) | 49 (5.9) |
| Average number of workflows used at moment of inclusion – n (range) | 1.5 (1–4) |
| Average number of experiments used at moment of inclusion – n (range) | 1.5 (1–6) |
| **Diagnostic Track** |  |
| Genetic testing prior to inclusion – n (%) | 106 (12.7) |
| Average number of requests – n (range) | 2.1 (1–8) |
| Average time spent in months – n (range) | 67 (1–317) |
| Genetic testing after inclusion – n (%) | 133 (16.0) |
| Average number of requests – n (range) | 1.4 (1–7) |
| Average time spent in months – n (range) | 7 (1–30) |

### Study design and SoC

All samples underwent two genetic testing pathways, consisting of SoC, followed by lrGS mimicking a prospective parallel design **(Figure 1A**). Diagnostic testing and reporting in SoC followed routine procedures under our ISO 15189-certified accreditation. Additionally, SoC testing data before (<2022) and after study inclusion (censored at 12-12-2024) were also collected for insight into the diagnostic tract.

#### lrGS

For all samples, an automated library preparation was performed, followed by PacBio HiFi lrGS using one Revio SMRT cell per sample, according to the manufacturer’s protocol, to an estimated coverage of 30x **(Table S2**). Data were processed using an in-house custom bioinformatics pipeline (**Figure S1**). Further details are provided in **Supplementary Appendix 1**.

### Variant interpretation

Variant interpretation was performed using the original SoC diagnostic request, containing the index case’s phenotype as well as the requested gene(s) or disease-gene panel(s). A custom graphical user interface facilitated variant prioritization mimicking SoC diagnostic interpretation strategies (**Supplementary Appendix 1**).

### Comparison of outcome measures between SoC and lrGS

SoC and lrGS were analyzed independently at the index case-level by separate teams, with results blinded between pathways. Each pathway yielded one of three outcomes: a conclusive, possible, or no genetic diagnosis (**Supplementary Appendix 1, Table S13**). Outcomes were then compared for concordance or discordance, with discordant cases investigated in detail.

### Impact analysis

Data from the direct comparison of SoC and lrGS were extrapolated to the centers’ combined annual germline testing in 2024 (n=15,150 index cases) to evaluate effects on diagnostic yield, including false negatives and newly identified diagnoses. These results were used to model scenarios assessing the impact of implementing a generic lrGS-based diagnostic workflow.

## Results

### Cohort demographics and diagnostic yield of SoC

The potential of lrGS as a generic diagnostic strategy was assessed by direct comparison with SoC in 832 index cases (84 trios, 748 singletons; 1,000 samples; **Figure 1A**). Cohort demographics (**Table 1**) were representative of annual diagnostic postnatal referrals for EDTA-based germline testing in our tertiary centers. At inclusion in 2022, a mean of 1.21 diagnostic requests per index case was submitted (**Table 1**; **Figure 1A**; **Table S1**), requiring a combination of molecular and cytogenetic workflows (n=12) and reanalysis of existing data.

On average, 1.5 experiments (range 1–5) using 1.5 workflows (range 1–4) were performed per index case. SoC yielded a conclusive diagnosis in 137 index cases (16.5%) and a possible diagnosis in 99 (11.9%), while 596 index cases (71.7%) remained undiagnosed (**Table 1**; **Figure 1B**; **Table S1**).

### Diagnostic yield of lrGS

To mimic an lrGS-first approach, we sequenced 1,000 genomes using HiFi lrGS at a median coverage of 29.2× and median read length of 14.7 kb (**Table S2**). Per genome, lrGS identified on average 5,335,455 SNVs/InDels, 67 CNVs, and 53,784 SVs (24,366 >50 bp in size; **Table S2**). Using variant prioritization and interpretation identical to SoC, lrGS yielded a conclusive diagnosis in 160 of 832 index cases (19.2%) and a possible diagnosis in 74 (8.9%), while 598 (71.9%) remained undiagnosed (**Figure 1B**).

### Diagnostic concordance between SoC and lrGS

We compared diagnostic outcomes at the index-case level to assess concordance between pathways. Overall concordance between SoC and lrGS was 96.4% (802/832; **Figure 1B**; **Tables S1, S3**). In 28 index cases (3.4%; **Figure 1BC**; **Tables S1, S3**), lrGS improved diagnostic outcomes. lrGS detected a previously missed InDel, CNV or SV (n=4), or refined CNV positioning (n=1) to upgrade a possible to a conclusive diagnosis (**Figure 2AB**). In the remaining 23 index cases, phasing information improved interpretation: 21 showed variants in *trans*, upgrading possible to conclusive diagnoses, while 2 showed variants in *cis*, excluding them as the cause of the phenotype. In the remaining two index cases (0.2%; **Figure 1BC**; **Tables S1, S3**), lrGS failed to detect pathogenic variants identified by SoC. The first was a low-grade mosaic SNV in *GJB2* (4% VAF) detected by SoC using targeted deep sequencing (4,511×); retrospective lrGS review showed the variant in 1 of 23 reads (**Figure S2**). The second was a *de novo* 1 bp deletion in *TLK2* identified by trio-based ES; lrGS analysis revealed low overall coverage (16×) with 11 reads at the locus, of which only two supporting the variant (**Figure S2**).

**Figure 2.**
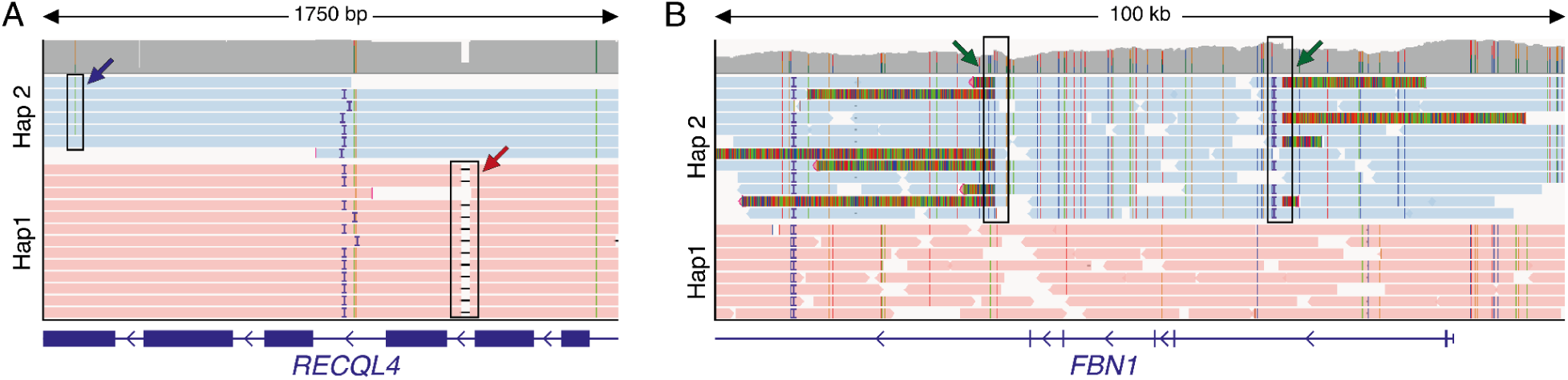
Showcases of lrGS-exclusive diagnoses. **A)** Aligned reads for IND_805 in region Chr8(GRCh38):g.144,512,823–144,514,673. Diagnosis was enabled by identifying and phasing a 24 bp intronic deletion in *RECQL4* that was missed in SoC (red arrow). The previously identified variant is in *trans* (blue arrow) (**Table S1; Table S3**). **B)** Aligned reads of IND_271 in region Chr15(GRCh38):g.48,557,346–48,567,346. Diagnosis was enabled by identifying and phasing an intragenic tandem duplication involving four exons of *FBN1* (breakpoints shown by green arrows) (**Table S1; Table S3**). Reads are grouped and colored by haplotag (red and blue), and InDels less than 6 bp are not shown.

### Longitudinal follow-up of genetic diagnostic referrals

The diagnostic journey for rare diseases is often lengthy, with few genetic diagnoses initially identified. We therefore analyzed the diagnostic track before, and after inclusion (**Table S1**). Of the 832 index cases, 106 (12.7%) had prior diagnostic testing (<2022) with no conclusive diagnosis. The average number of requests was 2.12 (range 1–8), starting 67 months before inclusion (range 1–317 months).

After inclusion, 133 index cases underwent additional diagnostic testing (mean, 1.41 requests; range, 1–7; **Table 1** and **Table S1**). Longitudinal follow-up of these cases within the SoC pathway identified 7 conclusive (5.3%) and 9 possible diagnoses (6.8%). Reanalysis of the corresponding lrGS data, mimicking the SoC workflow, yielded 6 conclusive (4.5%) and 10 possible diagnoses (7.5%).

Comparison of lrGS results to SoC showed 97.7% concordance (130/133). Two diagnoses (1.5%) were missed by lrGS, both involving GA-based repeat expansions, whereas one diagnosis (0.8%) was exclusively identified by lrGS (pathogenic SNV in a known disease gene; **Table S1**).

### Haplotype-based genomes for clinical interpretation

As phasing emerged as the main advantage of lrGS over SoC, we systematically evaluated its putative diagnostic impact. We assessed the ability to phase heterozygous SNVs/InDels within 4,887 autosomal rare disease genes (**Table S4**). Using HiFi lrGS, an average of 91.4% of heterozygous variant pairs within the same gene were phaseable (**Figure 3A**; **Table S4**). Phaseability correlated with gene length, reaching 98.7% for genes ≤39 kb, which comprises 50% of disease-associated genes (**Figure 3A**).

**Figure 3.**
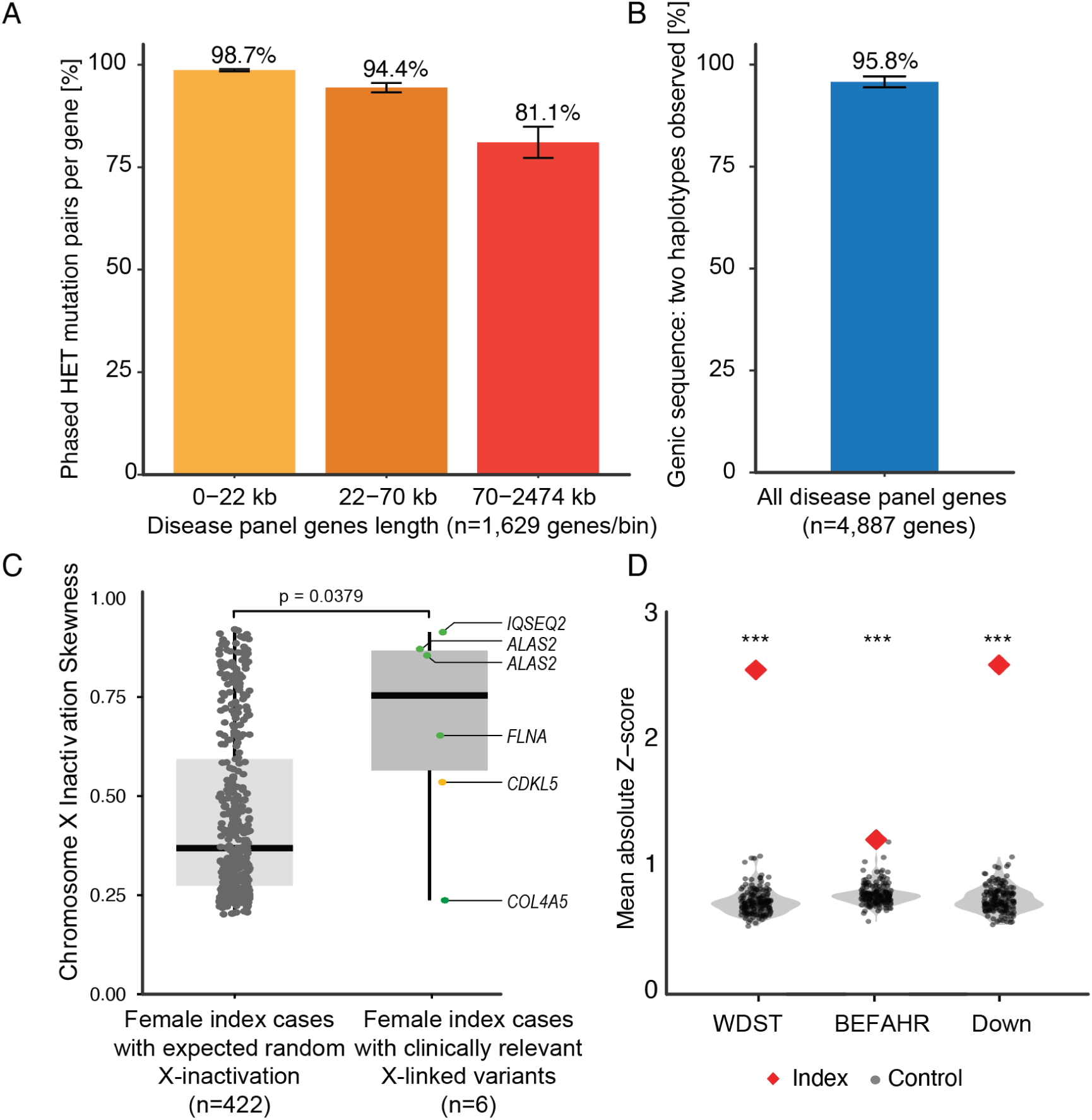
Haplotype phasing and methylation analysis. **A)** A histogram showing the percentage of modeled heterozygous (HET) variant pairs in the same disease panel gene that can be phase-resolved. Autosomal genes with a known disease association (n=4,887) are grouped into three equally sized bins (n=1,629) based on gene length (**Table S4**) **B)** Percentage of genic positions (i.e. including UTRs and introns) within disease-associated genes for which we find two distinct haplotypes. **C)** Analysis of skewed X-chromosome inactivation in female index cases. In female index cases with clinically relevant X-linked variants (right panel), a significant skewing of X-inactivation can be observed when compared with female index cases without clinically relevant X-linked variants (left panel) Green dots: (likely) pathogenic variants; orange dot: variant of unknown significance (**Table S5**). **D)** DNAm results for the three index cases for which the methylation profile was significantly different compared with controls (i.e. healthy parents of index cases, n=168). Index cases carried pathogenic variants in *KMT2A, TET3* and a trisomy of chromosome 21, causing Wiedemann-Steiner (WDST) syndrome, Beck-Fahrner (BEFAHR) syndrome, and Down syndrome, respectively. Each panel shows one DNAm analysis, with black dots and gray areas showing the distribution of lrGS-based 5mC mean absolute Z-scores in controls. Red diamonds mark the index case’s mean absolute Z-score. Higher scores indicate greater deviation from controls indicating aberrant 5mC profiles (**Table S6**). P-values were calculated using a one-sided normal test; ***p<0.001.

The phasing N50, i.e. the distance at which 50% of SNV pairs remain phaseable, was ∼200 kb for lrGS, compared with ∼0.8 kb for srGS (**Figure S3**). Phase-resolved haplotypes also enabled detection of allelic drop-out. In lrGS, 95.8% of positions across all 4,887 disease-associated genes were covered by phase-resolved reads, allowing exclusion of allelic drop-out at these loci (**Figure 3B**).

### Beyond genetic variation detection: epigenetic read-out

An additional advantage of lrGS is its ability to directly detect DNA methylation. We therefore evaluated the diagnostic utility of lrGS-derived 5mC data. Although no imprinting disorders were identified among the 832 index cases (**Figure S4**), lrGS revealed strongly skewed X-inactivation in three of six female cases carrying clinically relevant X-linked variants (**Figure 3C; Table S5**).

We next assessed the use of 5mC-based DNAm profiles. In 6/832 index cases, a (likely) pathogenic variant was identified in a gene with an available disease-specific DNAm profile. In three index cases, DNAm profile-specific CpGs showed significantly altered methylation compared with controls, confirming the disease-specific DNAm profile and providing additional support for variant pathogenicity (**Figure 3D**). In two index cases, lrGS did not confirm the expected DNAm profile, while in one index case only two CpGs remained after filtering, which was insufficient for analysis (**Supplementary Appendix 1; Table S6**).

### Operational, analytical and economic considerations of lrGS clinical adoption

Although lrGS is supported as a diagnostic test, additional operational factors need assessment before implementation, including the potential increase in interpretation workload due to higher variant yield. To assess this, we estimated variants requiring review in a simulated median-sized disease-gene panel (187 genes; **Supplementary Appendix 1**). Across 5,000 simulated panels, lrGS and ES each detected an average of 11 SNVs/InDels per panel, compared with 13 for srGS (**Figure S5**; **Table S7**). For SVs, lrGS identified 0.39 events per panel, versus 0.17 for ES and 0.34 for srGS, indicating no substantial increase in interpretation burden.

Prompted by variants missed by lrGS and known challenges in GA-rich regions, we analyzed systematic coverage dropouts. Among 4,887 autosomal disease-associated genes, 53 (1.1%) contained regions with <50% median coverage in >90% of samples, including 12 with severe dropouts (<25%). Most dropouts (71.7%) were attributable to GA-rich regions; others were linked to non-GA repeats (n=4), reference genome gaps in GRCh38 (n=5), or segmental duplications exceeding read length (n=4; **Figures S6–S9; Table S8**). Nonetheless, 29/51 genes still have >90% of the gene covered with >50% of median genome-wide coverage, suggesting that most variants in these genes can still be reliably detected (**Table S8**).

To assess cost-reduction potential, we evaluated the impact of reduced sequencing depth by bioinformatic down-sampling of the aligned reads of 156 index cases with a conclusive diagnosis (187 variants) from 30× to 20×, 15×, and 10× coverage. This resulted in variant recall reductions of 1.1%, 3.2%, and 7.8%, corresponding to loss of diagnosis in 1–2 cases at 20× and 12–13 cases at 10× (**Table S9**; **Figure S10**). SV recall rates (>50 bp) were disproportionately affected by low coverage (−3.6%, −10.0%, −15.4%; **Figure S10**).

### Modeling the diagnostic yield of a generic first-tier lrGS approach for rare diseases

We evaluated whether a generic lrGS-first strategy would improve rare disease diagnostics by modeling replacement of all SoC tests with a single lrGS assay for all index cases referred to our tertiary diagnostic centers in a single year. In 2024, 17,497 data-generating diagnostic requests were made for 15,150 index cases (mean 1.2 requests per index; **Figure S11**). Of these, 2,491 (16.4%) received a conclusive diagnosis and 1,311 (8.7%) a possible diagnosis, while 11,348 (74.9%) remained undiagnosed. Most diagnoses were due to SNVs/InDels (n=3,728, 78.8%), largely detected by ES and srGS (n=2,959, 79.1%; **Figure S11**).

We first modeled known lrGS limitations at 30× coverage, including detection of GA-based repeat expansions, low-level mosaicism (<15% VAF), heteroplasmic mtDNA variants (any level of heteroplasmy), and balanced events mediated by repeat exceeding HiFi read length. These were expected to result in missed diagnoses for 128 conclusive and 42 possible diagnoses (**Figure S12)**. Next, we modeled the positive impact of an lrGS-first approach on diagnostic yield using data from the parallel cohort (n=832) and large-scale lrGS studies, conservatively restricting extrapolation to genes with established disease-gene associations, and only restricted to the gene(s) from the SoC diagnostic request (**Table S10**). All 2,363 conclusive diagnoses (i.e. other than those missed due to technical limitations) were predicted to be identified by lrGS. Among the 1,311 possible diagnoses, we extrapolated i) variant phasing in recessive disease genes, while accounting for gene-specific phaseability, ii) detection of a second (likely) pathogenic variant in recessive genes, and iii) DNAm profiling. This shifted 278 possible diagnoses to conclusive (203 phased in *trans*, 65 second alleles, 10 DNAm-positive) and 20 to no diagnosis due to variants in *cis* (**Figure S12**). Finally, based on disease-specific yield increases reported in large lrGS studies and weighted to our referral spectrum (**Table S11**), an additional 223 previously unresolved cases were predicted to shift from no diagnosis to a conclusive diagnosis (**Figure S12**).

Collectively, a generic lrGS-first approach at 30× would improve diagnostic outcomes in 521/ 15,150 index cases (3.4%), while 170 cases (1.1%) would be identified only by SoC. This would significantly increase the conclusive diagnostic yield from 16.4% to 18.9% (two-tailed Fisher’s exact test, p < 0.0001; **Figure 4**). Reducing coverage to 20× would result in only a marginal loss of clinically relevant findings, given the 99.6% recall rate for SNVs/InDels (**Table S12**).

**Figure 4:**
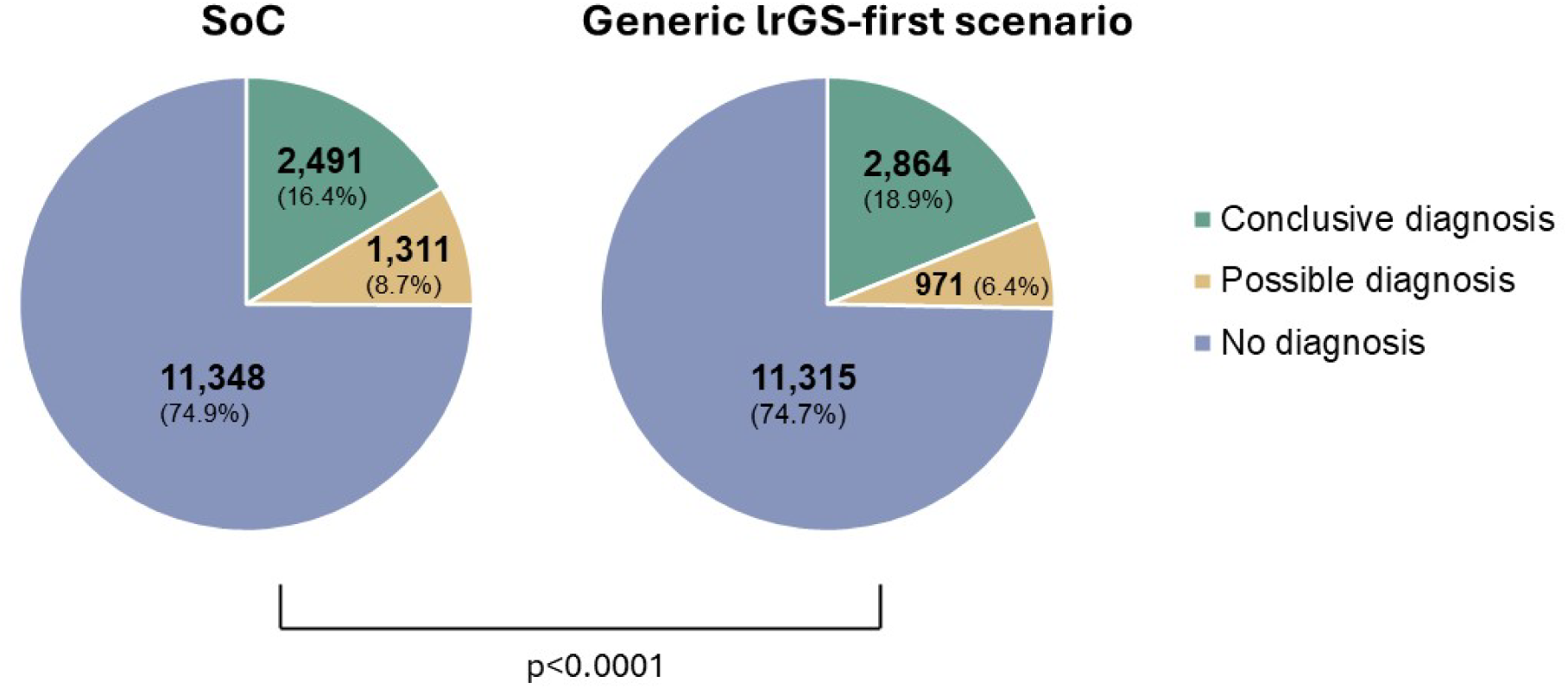
Predicted diagnostic yield for generic lrGS-first approach for germline-based testing for rare diseases. In a model-based analysis, lrGS advances for variant interpretation and improved variant detection (limited by known disease-gene associations, and gene(s) from the original SoC request) were considered, alongside technological limitations leading to missed diagnoses (**Figure S12**). The overall diagnostic yield for index cases referred for germline-based testing in 2024 (n=15,150) would significantly increase from 16.4% in SoC to 18.9% in lrGS (p<0.0001, two-sided Fisher’s exact test).

## Discussion

Over the past decade, srGS has increasingly been adopted as a first-tier diagnostic test for rare diseases, supported by clinical utility studies^6,17^. Its value lies in access to the non-coding genome and broad variant detection in a single assay. However, technical limitations in complex genomic regions, such as highly homologous sequences and repeat expansions, have prevented srGS from fully replacing complementary cytogenetic and molecular workflows^7^. Recent studies have shown that lrGS overcomes these limitations and achieves accuracy comparable to short-read technologies^16^,. Here, we demonstrate that lrGS can serve as a generic first-tier test for germline diagnostics using two complementary approaches. First, direct comparison with SoC (n=832) showed 96.4% concordance, with lrGS outperforming SoC in 3.4% of cases and SoC outperforming lrGS in 0.2%. Second, modeling a generic lrGS-first strategy for annual diagnostic referrals predicted a 2.5% increase in diagnostic yield while replacing all other workflows. Together, these findings support lrGS as a generic first-tier test that improves diagnostic yield and laboratory efficiency.

While lrGS appears to be the most comprehensive first-tier test for rare disease diagnostics, several detection challenges remain, which are non-unique to lrGS, including GA-rich regions, low-level mosaicism including mtDNA heteroplasmy and balanced SVs mediated by repeats exceeding the read length. In our model this translated into 170 false negative results. Reduced performance of lrGS in GA-rich regions has been previously reported^18,19^ and reflects polymerase-related sequencing bias. Detection of long GA-rich alleles improves with higher sequencing coverage, achievable through inclusion of non-HiFi reads^16^ or by using CRISPR-based targeted assays^20^. Although the latter adds an auxiliary test, most GA-based repeat expansions arise from a small number of loci (*RFC1* [MIM 614575], *FGF14* [MIM 193003 and MIM 620174], *FXN* [MIM 229300]) associated with recognizable ataxia phenotypes, making targeted follow-up testing feasible. Detection of low-level mosaicism is coverage-dependent. While increased coverage can improve detection of nuclear mosaicism, mtDNA heteroplasmy remains challenging due to underrepresentation of the 16-kb circular mtDNA in lrGS libraries. In our cohort of 832 index cases, all clinically relevant mtDNA variants were detected, including two homoplasmic and one 17% heteroplasmic. For modeling, however, we conservatively assumed heteroplasmic mtDNA variants would be missed in an lrGS-first strategy, although detection of a subset is plausible. As an alternative, targeted mtDNA sequencing remains an effective adjunct in selected cases.

The generic nature of lrGS allows consolidation of cytogenetic and molecular workflows, reducing or avoiding iterative testing, thereby reducing time to diagnosis. This benefit has been quantified in a recent comparison of SoC testing versus a single test lrGS approach^21^. Whereas that study focused on direct comparison of the initial request, we extended this observation using longitudinal data of the 832 index cases, showing that 24.2% received either prior or post-inclusion testing. In the modeling cohort, the mean number of diagnostic requests per index case was already 1.2, with 2,212 cases (14.6%) receiving multiple requests within the same year. Beyond inability of the initial test to establish a diagnosis (i.e. “the diagnostic odyssey”), repeated testing most often reflected the need to further characterize variants using alternative methods or to identify variants potentially missed by the initial assay, information that could, in principle, be obtained from a single lrGS analysis.

Phasing information provides substantial clinical value. In recessive disorders, as demonstrated by our data, it establishes whether pathogenic variants are in *trans* or in *cis*, confirming disease causality without the need for additional family samples. This is particularly valuable for index cases without available familial samples, reduces the time of diagnostic uncertainty when segregation testing is required and decreases healthcare resource use. Furthermore, for repeat disorders, lrGS enables precise allele-specific repeat sizing, including motif composition and interruptions. Finally, comprehensive variant detection facilitates interpretation of variants in the context of all detected alleles, consistent with a Bayesian approach to genomic analysis informed by allele-specific locus assignment.

Although this study used lrGS exclusively to mimic SoC requests, rather than attempting to maximize the diagnostic yield achievable with lrGS, we nevertheless predicted an absolute increase of 2.5% in diagnostic yield, reflecting 15% relative increase over current practice. This gain primarily derives from phasing, incorporation of 5mC data, and improved detection of variant types not detectable in the SoC data. For index cases without an initial genetic diagnosis, lrGS data can be retained for future reinterpretation, offering a superior substrate compared with srGS because of its greater completeness and near haplotype-resolved genome representation^22,23^.

Implementation of a generic lrGS workflow depends on multiple laboratory-specific factors, including lab size, number of SoC workflows, level of automation, and costs associated with the transition to, and routine operation of, lrGS. Our study focused on quantifiable operational and analytical aspects of postnatal germline diagnosis of rare diseases, for which we observed no major hurdles in technical or clinical validation or in data interpretation. In our laboratory, most samples are extracted from EDTA whole blood. Although similar performance is expected for other DNA sources (e.g., buccal smear, skin biopsy, or muscle biopsy), this would require verification. Likewise, additional optimization may be needed for prenatal DNA sources, such as chorionic villi or amniocytes.

From an economic perspective, we limited our analysis to the impact of reducing lrGS coverage on recall rates of (likely) pathogenic variants. A full micro-costing assessment of a generic lrGS approach would need to balance potential cost reductions from phasing out existing workflows and workforce changes against possible increases in per-sample sequencing costs^24,25^, for which insights are currently not yet available. At present, data production costs for lrGS remain higher than for srGS. However, recent announcements promise cost reductions, with a reported price range of $300-$345 per genome for Oxford Nanopore Technologies and PacBio. Although our study evaluated only HiFi sequencing, our main conclusions are expected to be generalizable to nanopore sequencing and reflect the advantages of lrGS rather than platform-specific effects.

In summary, our study provides evidence for clinical utility of lrGS as a first-tier, generic assay for germline genetic testing. Whereas routine implementation may vary across diagnostic laboratories, our data support a more comprehensive assessment of individual clinical genomes and the feasibility of large-scale deployment in health care systems.

## Data Availability

Our study is performed on sensitive patient data, which are subject to institutional review board restrictions. We are thus unable to submit primary genotype data to a public database. All data available for sharing, particularly information about pathogenic variants, are contained in the manuscript and supplementary appendices. We are open to legitimate data requests, which will be process according to national and institutional guidelines.

## Acknowledgements/Funding

We thank our colleagues from the divisions of Clinical Genetics and Genome Diagnostics of the Department of Human Genetics, Radboudumc and Clinical Genetics of Maastricht UMC+, as well as the Radboud Genome Technology Center for technical support.

AH was supported by a ZonMW (The Netherlands Organization for Health Research and Development) Vici grant (No. 09150182310053). LS was supported by a Sigrid Jusélius Foundation fellowship (220540). The project received funding (to TJJDB, AH and LELMV) from the Dutch Ministry of Economic Affairs by means of a PPP Allowance made available by the Top Sector Life Sciences & Health to stimulate public-private partnerships. The aims of this study contribute to the Solve-RD project (to MS, AH, CG and LELMV, grant agreement N°779257), ERDERA project (to BVDS, LS, MDG, MS, AH, CG and LELMV, grant agreement N°220540), and a Marie Sklodowska-Curie project (to WH, grant agreement N°101150006) which received funding from the EU Horizon 2020 and EU Horizon Europe research and innovation programs. Views and opinions expressed are those of the authors only and do not necessarily reflect those of the financing bodies or any other granting authority, who cannot be held responsible for them.

The authors of this publication are members of the European Reference Networks ITHACA (LSB; CG; LELMV), RITA (AH), RND (EJK), and GENTURIS (ARM, MJLL, WAGVZS) EU grant N°101156387).

## Conflict of Interest

AH, CG and LELMV have received travel support from Pacific Biosciences of California, Inc. AH has received travel support from Bionano Genomics, Inc. Pacific Biosciences of California, Inc. also kindly provided part of the reagents required for this study. The remaining authors declare that they have no competing interests. QS is a shareholder in Pacific Biosciences of California, Inc., Oxford Nanopore Technologies plc., and Illumina, Inc..

## Notes

### Author Declarations

The study confirmed the principles of the Helsinki Declaration. In addition, the study was performed as part of a local validation study for the implementation of GS under ISO15189 accreditation and assessed as a diagnostic innovation by the Medical Ethics Review Committee Arnhem-Nijmegen under dossier number 2020-7142

