## Supplementary Appendix 1 for "Clinical long-read genome sequencing for rare disease diagnostics"

Supplement to: Clinical long-read genome sequencing for rare disease diagnostics

Methods ... 3

Cohort selection and representativeness ... 3

Long-read sequencing & variant interpretation ... 4

Automated library preparation and HiFi sequencing ... 4

Bioinformatic data analysis ... 4

Variant prioritization, interpretation, and classification ... 5

Comparison of diagnostic outcome measures between SoC and lrGS ... 5

Assessing quality parameters of generic lrGS approach ... 6

Phasing analysis ... 6

Phasing analysis per gene ... 6

Identification of low-coverage genomic regions ... 6

Coverage near GA-rich regions ... 6

Variant recall in titration experiments ... 7

Methylation-based analysis ... 8

Imprinting analysis ... 8

Skewed X-inactivation analysis ... 8

DNA methylation profile analysis ... 8

Diagnostic implications of a generic lrGS approach ... 9

Interpretation burden ... 9

Impact analysis on diagnostic outcome ... 9

Figures ... 11

Figure S1: Bioinformatic pipeline for aligning, variant calling and annotation ... 11

Figure S2: Variants missed with lrGS ... 12

Figure S3: Phasing in lrGS and srGS ... 15

Figure S4: Visualization of 5mC data in IGV for four index cases tested for Prader-Willi and Angelman imprinting disorders. ... 16

Figure S5: Number of variants for interpretation after routine diagnostic filtering steps. ... 17

Figure S6: Genes with impaired coverage ... 19

#### Methods

##### Cohort selection and representativeness

The Department of Human Genetics of the Radboudumc and the Department of Clinical Genetics of Maastricht UMC+ are tertiary referral centers for genetic diagnostic testing. For the purpose of this study, we evaluated all 24,570 index cases referred for germline-based diagnostic testing to find the genetic cause underlying their rare disease in 2022 in both centers. This included n=30,514 diagnostic requests, across 883 different diagnostic indications[1]. We used this overview as a start to generate a comprehensive selection of 1,000 samples to be representative for these medical centers' joint annual germline-based clinical testing for the primary diagnosis of germline variants underlying hereditary and congenital diseases using blood-derived DNA. Additionally, this cohort was representative in terms of contribution to the yearly diagnostic requests, diagnostic yield, and standard of care (SoC) workflows used.

First, we only included the first diagnostic referral for each index case under the assumption that all further referrals would include a re-analysis of the existing (long-read) data. In addition, we only focused on diagnostic indications for which a germline variant was expected (e.g., excluding those related to somatic variant detection, i.e., hematological malignancies and solids cancers). This left 592/883 diagnostic indications, each requested between 1 and 982 times in 2022 (median 4, IQR 1-18, with the top 20 most frequently requested indications responsible for 48.2% of all requests).

To verify that our selection was still representative for our current production, we extracted all diagnostic referrals between June 1st and June 30th of 2023 and evaluated the relative frequency of each indication for comparison to the 2022 data. Differences observed were manually checked and curated. These included renaming of (the same) diagnostic indications, diagnostic indications for which no longer testing is indicated in our laboratories, and diagnostic indications that were not requested in 2022 but were in 2023. In addition, we corrected for the number of single-gene requests (often requested ≤1 time/year) to avoid unequal representation of those indications, leaving 322 diagnostic indications.

Next, we determined the relative contribution of these 322 indications to calculate how many samples should be included per indication in a series of 1,000 samples to match the relative contribution of this clinical entity to our yearly overall referrals. Given the unequal distribution of requests per diagnostic indication, we set a maximum number of samples to sequence per indication at 50 index samples. In addition, for 34 diagnostic indications trio-based analysis was requested, requiring 3 samples per request. For trios, we set a maximum of 30 index cases per indication (i.e. 90 samples per indication) to maximize the inclusion of different diagnostic indications. This selection resulted in 832 index patients covering 322 different diagnostic indications.

For all index cases, we subsequently collected information on genetic diagnostic testing prior to inclusion as well as post inclusion (censored until 12-12-2024), including for each request: the date of request, the diagnostic outcome, cytogenetic and/or molecular workflow(s) used, and clinically reported variants.

The study confirmed the principles of the Helsinki Declaration. In addition, the study was performed as part of a local validation study for the implementation of GS under ISO15189 accreditation and assessed as a diagnostic innovation by the Medical Ethics Review Committee Arnhem-Nijmegen under dossier number 2020–7142.

#### Long-read sequencing & variant interpretation

<u>Automated library preparation and HiFi sequencing</u>

For the long-read sequencing (lrGS), we performed a fully automated library prep module and annealing, binding, and cleanup (ABC) module using an NGS STARV MOA system from Hamilton Robotics. In brief, 3.3 µg high-quality genomic DNA per sample was used as input material and processed on the robot using the Revio HiFi Prep Kit 96 (PacBio, Menlo Park, CA, USA) according to the manufacturer's protocol. DNA was sheared to a target size of 15-18 kb using pipet tip shearing, and short fragments were removed using PacBio's Short Read Eliminator (SRE) kit. Concentration measurements were performed on-deck using Fluoreye in combination with Quant-it HS kit (Thermo Fisher Scientific, Waltham, MA, USA). Concentrations after SRE and prior to shearing were measured manually with Qubit dsDNA HS (Thermo Fisher Scientific) and samples with high concentrations were diluted accordingly before shearing. All samples were processed in batches of 24 samples, and each of the 24 samples was loaded on its own Revio SMRT cell (PacBio) as specified by the manufacturer. All sequencing was performed between August 2024 - January 2025, utilizing 3–4 Revio systems (PacBio). SMRT cells were sequenced to obtain 24-hour movies.

<u>Bioinformatic data analysis</u>

Samples were analyzed using an inhouse bioinformatic pipeline or long-read sequencing data as described before[2] with minor modifications, providing a semi-automated pipeline integrating different PacBio and in-house software tools with in-house annotation (**Figure S1**). In brief, high-fidelity (HiFi) reads were aligned against the GRCh38/hg38 reference genome using pbmm2 (v1.13.1). Minimal threshold for variant calling and annotation was set at 15x median coverage per sample, requiring resequencing for 7/1,000 samples (0.7%). Structural variants (SVs; pbsv v2.9.0) and small variants (DeepVariant v1.6.1) were called with phasing information (HiPhase v0.10.1) and annotated using publicly available databases. A specific analysis for copy number variants (CNVs) was performed using HifiCNV (v1.0.0) by detecting variation in depth of coverage. Repeats were called using TRGT (v1.0.0), visualized by TRVZ (v1.0.0), and finally annotated using an in-house pipeline. Variant calling for specific paralogs and pseudogenes were called using Paraphase (v.3.1.1). Methylation calls were generated using pb-cpg-tools (v.2.3.2).

Variant prioritization, interpretation, and classification

The annotated variant calls resulting from the secondary analysis pipeline were interpreted using a custom graphical user interface (GUI). Interpreters had access to the original test request form from SoC, but not to the results thereof. SNVs/InDels, CNVs, SVs, and repeat expansions were filtered in the same two-step process. For single-gene requests, only variants affecting the requested gene were considered (coding sequence +/- 8 bp of splice site region and CNVs + SVs affecting the gene, **Table S13**). For panels and open exome requests, a disease panel or exome filter, respectively, was first applied, together with control population allele frequency filters. The resulting variants were prioritized based on the variant's impact on biological gene function, disease mechanism, expected inheritance pattern, and interpreted in the clinical context of the patient (**Table S13**). For classification of small variants, we used the guidelines established by the Association for Clinical Genetic Science (ACGS) and the Dutch Society of Clinical Laboratory Geneticists (VKGL)[3], using a 5-class system (Class 1 to Class 5) similar to the American College of Medical Genetics and Genomics (ACMG/AMP)[4], whereas for structural variation and numerical abnormalities the European guidelines for constitutional cytogenomic analysis (Class 1 to Class 5) was used[5].

#### Comparison of diagnostic outcome measures between SoC and lrGS

The diagnostic outcomes in SoC as well as for lrGS was determined at the individual patient level (**Table S13**) and summarized to be one of the following:

1) No diagnosis. No obvious pathogenic variant(s) (of either variant type) were detected that could potentially explain disease.
2) Possible diagnosis. A variant(s) of unknown significance was identified in a previously established disease gene that could explain the patient's phenotype. Or alternatively, a pathogenic variant(s) in a candidate disease-gene(s) was identified with a potential relationship to (part of) the patient's phenotype.
3) Conclusive diagnosis. A (likely) pathogenic variant in a disease gene associated with the patient's phenotype was detected, which explains the phenotype observed.

To objectively compare the diagnostic outcomes of both SoC and lrGS, diagnostic endpoints of lrGS and SoC were only disclosed for comparison after completion of the entire workflow, and comparison was performed by a single, dedicated assessor, who was kept blinded to the individual tracks. Discrepant diagnostic outcomes were reviewed by a multidisciplinary team including molecular geneticists, clinical laboratory geneticists, and clinical geneticists.

**Assessing quality parameters of generic lrGS approach**

<u>Phasing analysis</u>

We devised a test to estimate the probabilities for a hypothetical pair of SNVs that would be phased in any given sample. For each sample, we extracted the start- and end- positions of each phase block from the phased SNP VCF file produced by HiPhase (version 1.4.2)[6]. We then simulated 1,000 pairs of SNVs of a given distance (500 bp, 1–10 kb (in increments of 1 kb), 10-100 kb (in increments of 10 kb), 100 kb–1 Mbp (in increments of 100 kb), 2 Mb) and tested whether they would both fall into one read length distance (15 kb) within the same phase block, meaning that these SNPs would be phased if they actually occurred. The same was repeated for each sample, each time taking the real phase-blocks.

<u>Phasing analysis per gene</u>

We also estimated the fraction of variants that can be phased per gene as follows. Two variants were considered phaseable if they were within one read length (15 bp) or both located inside the same phase block. We reasoned that any SNV pair within a gene could be represented as coordinates on a 2D plane. In this framework, phaseable pairs would fall within a diagonal band (single-read connections) or within square regions (existing phase blocks). The combined area of these regions, relative to the total, defines the estimated co-phasing probability (**Figure S13**).

<u>Identification of low-coverage genomic regions</u>

We used Mosdepth[7] (version 0.3.8) to generate genome-wide coverage statistics for all samples in bins of 2 kb windows. For each sample, we next calculated a genome-wide median coverage (excluding ChrX/Y and all centromeres) and identified bins with <50% and <25% of the sample's median coverage. We then aggregated these annotations across all samples, identifying bins which were consistent (at least 90% of samples) in displaying reduced coverage. These 'systematically reduced' bins were then intersected with genome annotations of 4,887 autosomal disease genes to identify genes overlapping with at least one systematic low-coverage bin (<50% coverage compared to median), and with a very low-coverage bin (<25%).

<u>Coverage near GA-rich regions</u>

To test for the effect of GA (or reverse complement CT) -rich simple repeats on the read coverage, simple repeats on hg38 based on the Repeat Masker tool were obtained through the University of California Santa Cruz (UCSC) data portal[8]. Only repeats with more than 90% GA (or the reverse complement CT) content were retained. GA-rich regions which were overlapping or closer than <100 kb to other repeats were filtered to avoid measuring overlapping coverage effects. We grouped the GA-rich regions by length and measured mean coverages across all samples around these repeats using Mosdepth (version 0.3.8) (**Figure S8**).

Variant recall in titration experiments

We tested for the relation of read depth and variant recall by creating and analyzing datasets which were artificially downsampled to target coverages of 10x, 15x, and 20x using a procedure described previously[2]. Aligned read files (.bam) from all samples were subsampled to target coverages of 10x, 15x and 20x using the "*samtools view -b"* command (samtools version 1.21). This process was repeated 10-times for each target coverage, resulting in a total of 30 subsampled readsets per sample, or 10 per target coverage. Downstream processing and variant calling were done analogously to the original datasets, resulting in independent variant callsets for each permutation. Three samples with a coverage of less than 20x were excluded from this analysis.

In the original dataset, these samples together carried 187 pathogenic variants which could be automatically detected using at least one of the variant callers. Due to cross-calling between different tools (e.g., HiFiCNV and pbsv both calling certain CNVs), those 187 variants were represented by 201 individual variant calls. We checked for recall of these 201 calls in the subsampled datasets with a script implementing the following criteria (https://github.com/WHops/lrs100_downsample) (**Table S9**).

- SNVs: Same substitution within ±1bp.
- Mitochondrial SNVs additionally: Affected fraction within 20 percent points of original call (e.g. using original VAF as proxy for heteroplasmy).
- InDels (≤50 bp): All breakpoints within ±10 bp, and event length within ±5 bp of the original call.
- SVs (>50 bp, excl. translocations): The re-call and original call share ≥50% reciprocal overlap.
- Translocations: All breakpoints found within ±50bp of the original call.
- Repeats: Within ±1 kb: TRGT motif copy number on both alleles identified (±20% CN deviation allowed).
- Gene/Pseudogene CNVs (Paraphase): The total copy number of genes + their pseudogenes are reported correctly.

Of note, HiFiCNV sometimes reports 'split' calls, *i.e.* multiple consecutive calls that cover a single actual event. To alleviate this potential discrepancy, all neighboring CNV calls were always merged if they were at most 1 kb apart.

For repeat expansions, we tested for the motif and its repeat count on both alleles and allowed for plus/minus 20% in both. In cases where the repeat count of the motif was denoted as '>n copy numbers', we only tested for that allele to have at least a copy number of n in each given case. Source 'para_json' denotes variants that are exclusively called in the *.json files created by Paraphase. These are copy number changes affecting gene/pseudogene combinations. In the rare case that a copy-number change was correctly noted, but ascribed to the wrong gene copy, we accepted that paraphrase had ‘in principle’ identified the variant and counted this variant as recovered.

#### Methylation-based analysis

Imprinting analysis

We assessed which of the 832 index cases were referred for testing of imprinting disorders during the diagnostic track. This yielded four index cases, including referrals for Prader-Willi syndrome (PWS, n=3) and Angelman syndrome (AS, n=1). For these four cases, which all received a negative SoC result, we manually inspected reads across the *SNRPN/SNURF* promoter for allele-specific methylation and coverage differences, using the Integrative Genomics Viewer (IGV).

Skewed X-inactivation analysis

Haplotype-specific methylation data were generated using CpG Tools[9]. CpG sites for ascertaining Chromosome X-inactivation were selected based on multiple criteria: intersecting a CpG island (UCSC CpG islands on ChrX[8], methylation signal in at least half the male samples (n=240) and a mean methylation signal <5 in males. To exclude imprinted regions, only sites measured in at least half the female cohort (n=253) with a mean methylation score >25 were used. This resulted in 14,891 Chromosome X inactivation-relevant CpG regions.

For these sites, the haplotype methylation delta was calculated per sample using the following formula with hap1 and hap2 being the respective haplotype-specific methylation scores of a site:

$$\Delta haplotype\ methylation = |hap1 - hap2| \,/\, (hap1 + hap2)$$

The median haplotype methylation delta value per sample was used as a metric of Chromosome X inactivation skewing. Median haplotype methylation values were compared between patients carrying autosomal variants (n=105) and those with variants on Chromosome X (n=6) using a pairwise Wilcoxon rank-sum test.

DNA methylation profile analysis

To assess whether DNA methylation (DNAm) profiles could be detected in lrGS, we first made an overlap between publicly available DNAm profiles[10,11] and the reported variants in our 832 index cases. For index cases with a (likely) pathogenic variant in a gene with a published methylation profile, we used the corresponding set of CpG probes reported in the literature. To improve signal detection from 30x genomes, we included only probes with a reported case-control methylation difference greater than 15%. The lrGS methylation values in the index for each of the remaining probes were compared to those of controls. This control set consisted of the 168 unaffected parents (84 females and 84 males) that were included in this study for the purpose of trio-based analyses. For each CpG site, we calculated the mean and standard deviation in the control group to define a reference distribution. Z-scores were then computed for each probe in the sample, and the mean absolute Z-score was used to summarize deviation from the control. A one-sided

Z-test was used to assess whether the sample showed significant deviation, indicating presence of the DNAm profile.

#### Diagnostic implications of a generic lrGS approach

Interpretation burden

To model the expected interpretation burden of lrGS, we first generated 5,000 simulated gene panels, each containing 187 genes. The target size of 187 genes was chosen because it represents the median number of genes in all in-house diagnostic disease-gene panels used in our laboratories. The simulated panels were constructed from the full set of 5,166 known disease-associated protein-coding genes for which genetic testing was requested for at least one of the 832 index cases (**Table S14**)[12]. For each panel, 187 unique genes were selected using simple random sampling without replacement. Sampling was performed independently for each of the 5,000 iterations, allowing genes to appear in multiple panels across iterations but ensuring that each gene appeared at most once within a single panel. A fixed random seed was used to ensure reproducibility.

Subsequently, for each of the 5,000 gene panels, we quantified the number of rare, coding SNVs, InDels, CNVs, and SVs for each of the 832 index cases. Here, rare refers to variants with an allele frequency of less than 1% in our in-house databases and gnomAD v4.1.0[13]. Only coding variants were considered, with CNVs and SVs required to overlap at least one coding region. CNVs were combined with SVs to obtain the total number of SVs per sample. Importantly, post-filtering variants were neither stored nor analyzed individually; only their total counts were retained. To compare lrGS with current first-tier genetic testing approaches, we analyzed 100 ethnically matched in-house short-read exome sequencing (ES) and short-read genome sequencing (srGS) datasets. For each dataset, we determined the number of rare SNVs, InDels and SVs remaining after filtering variants within the same 5,000 disease gene panels. Again, rare refers to variants with an allele frequency of less than 1% in our in-house databases and gnomAD, and only coding variants were considered. For srGS, CNVs, and SVs were again combined into a single SV count, whereas for ES, in the absence of an SV caller, CNVs were used directly in subsequent analyses. This resulted in a vector of 5,000 mean variant counts for each variant type (i.e. SNVs/InDels and SVs) and each method, one value per artificial panel (**Table S7**).

Impact analysis on diagnostic outcome

We modeled a scenario of the overall impact of a generic lrGS-based diagnostic workflow on our medical centers' joint annual germline-based clinical testing in 2024. All 55,150 diagnostic requests were evaluated for eligibility in the model. First, 31,128 requests were excluded because they concerned cascade screening (n=8,899) or were outside the current scope of lrGS replacement, such as non-high-molecular weight DNA-based and/or biochemical assays (n=22,229). Subsequently, requests involving non-index cases (n=5,431), required solely for the interpreting index case data, were excluded. This yielded 18,591 diagnostic requests corresponding to 15,686 index cases. Among these index cases, 14,659 were referred for at least

1 data-generating diagnostic request (e.g. requiring a wet-lab cytogenetic or molecular workflow with or without bioinformatic processing), another 491 were referred for both ≥1 data-generating request and ≥1 request for reanalysis of existing data (e.g. no wet-lab experiments, only bioinformatic processing) and 536 index cases were only referred for reanalysis of existing data.

Considering only the data-generating diagnostic requests, in total 15,150 index cases accounted for 17,497 such requests, which collectively required 20,045 data-generating experiments (**Figure S11**). For each of those 17,497 requests, we evaluated the diagnostic outcome, categorized the SoC outcomes as a 'conclusive', 'possible' or 'no genetic' diagnosis (see '**Comparison of diagnostic outcome measures between SoC and lrGS'**). For all conclusive and possible diagnosis, variant(s) and variant type(s) were assessed and categorized per diagnostic workflow. Next, we modeled the outcome under a generic lrGS-approach, taking into account the (technical) limitations observed at 30x coverage, as well as the observed benefits from our series of 1,000 sequenced samples (**Table S10**) and supplement with observations from large-scale publications reporting on the diagnostic value of lrGS in specific clinical indications.

### Figures

##### Figure S1: Bioinformatic pipeline for aligning, variant calling and annotation

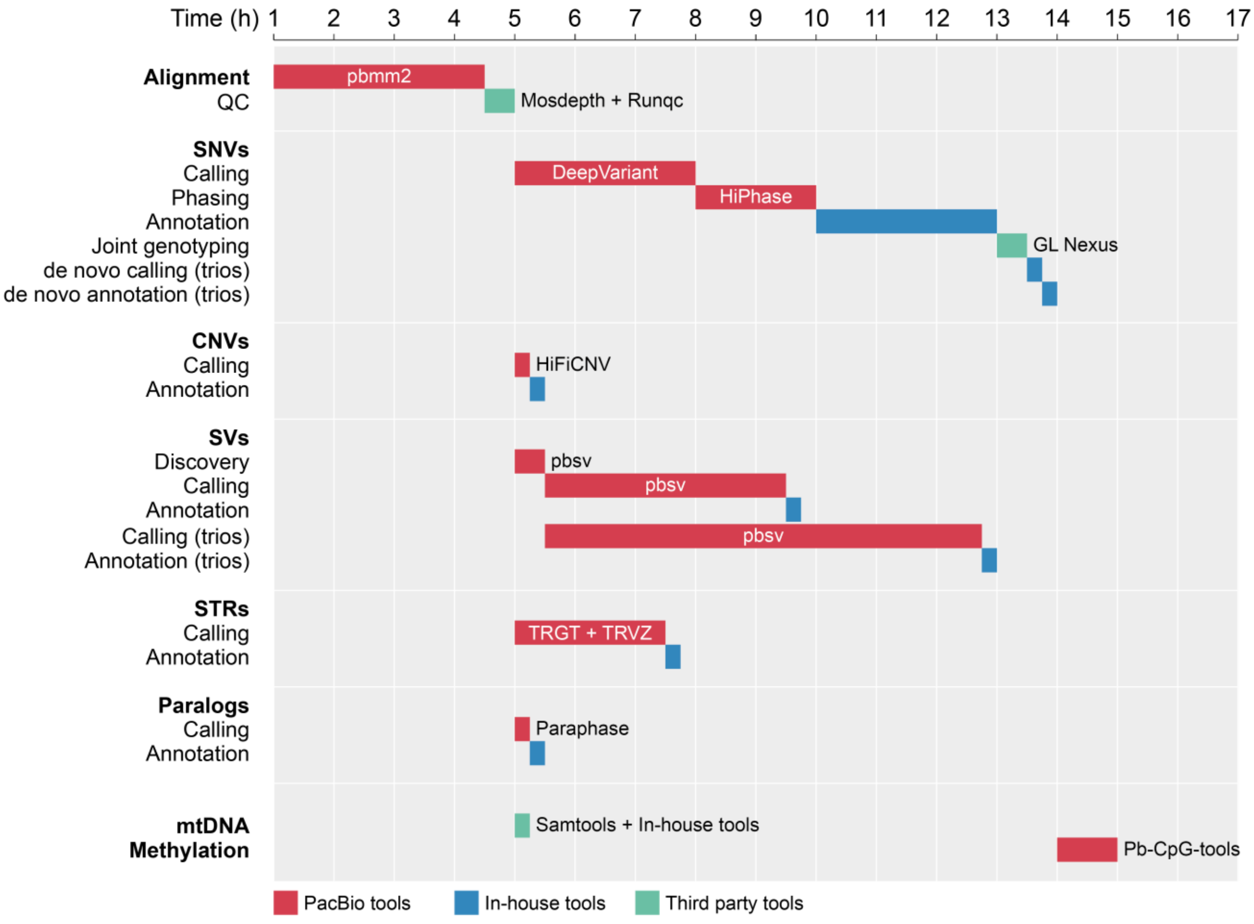

After read mapping (5 hours), each variant type was called by independent callers which were performed in parallel (10 hours), leading to an optimized pipeline of 15 hours from read mapping to variant annotation.

#### Figure S2: Variants missed with lrGS

A

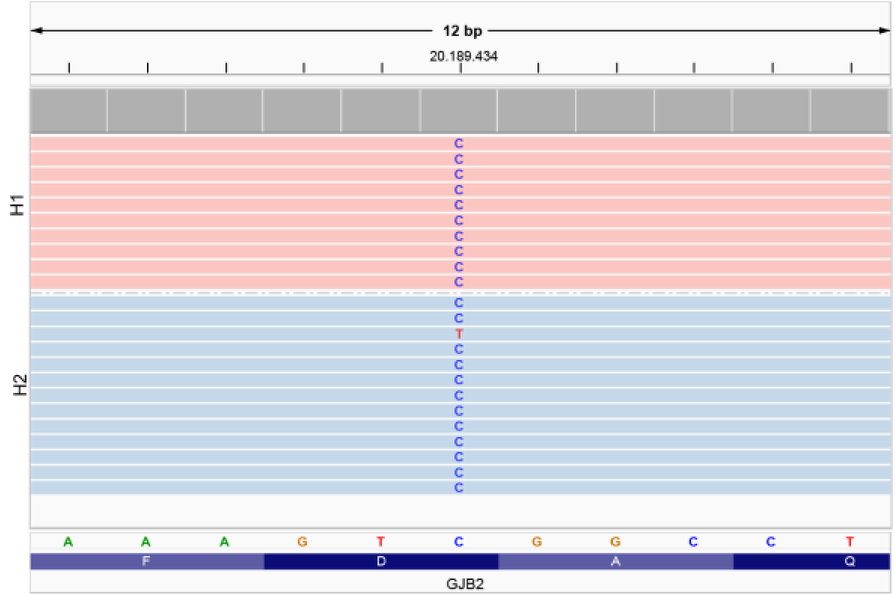

B

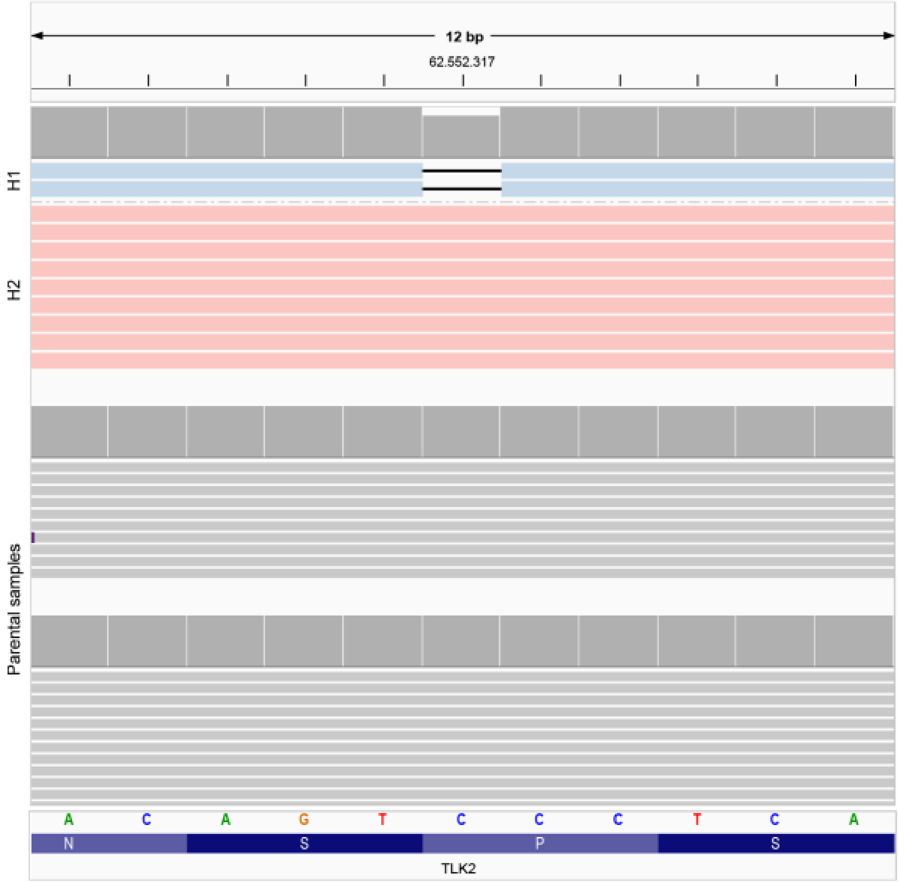

C

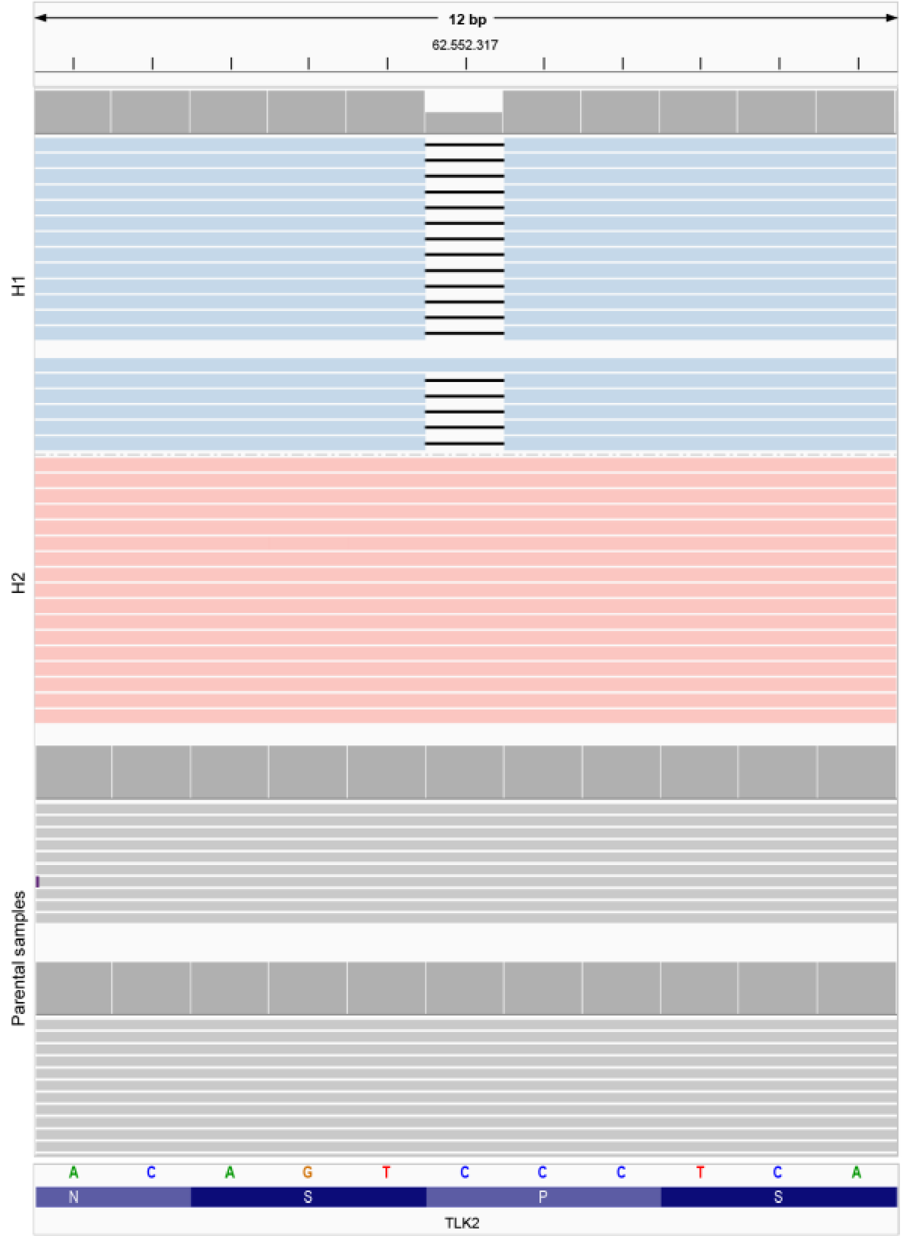

In two index cases (0.2% of cohort), lrGS failed to detect clinically relevant variants that were identified in SoC. **A)** The first case (IND_999) involved a low-level mosaic variant in *GJB2* that was detected in SoC using a targeted panel-based NGS approach. **B)** The second case (IND_36, parental samples IND_37 and IND_38) involved a *de novo* single base deletion in *TLK2*. This index sample achieved only 16x genome-wide coverage in lrGS, resulting in only 2/11 reads (18.2% VAF) containing the variant. Whereas this variant was called by the pipeline, it did not pass routine filtering thresholds, resulting in a false-negative lrGS outcome. **C)** Upon repeating the lrGS of the index, sufficient coverage was obtained, which led to the detection variant as expected. Note that the IGV screen shots presented correspond to the IrGS results, and not to the original SoC results.

**Figure S3: Phasing in lrGS and srGS**

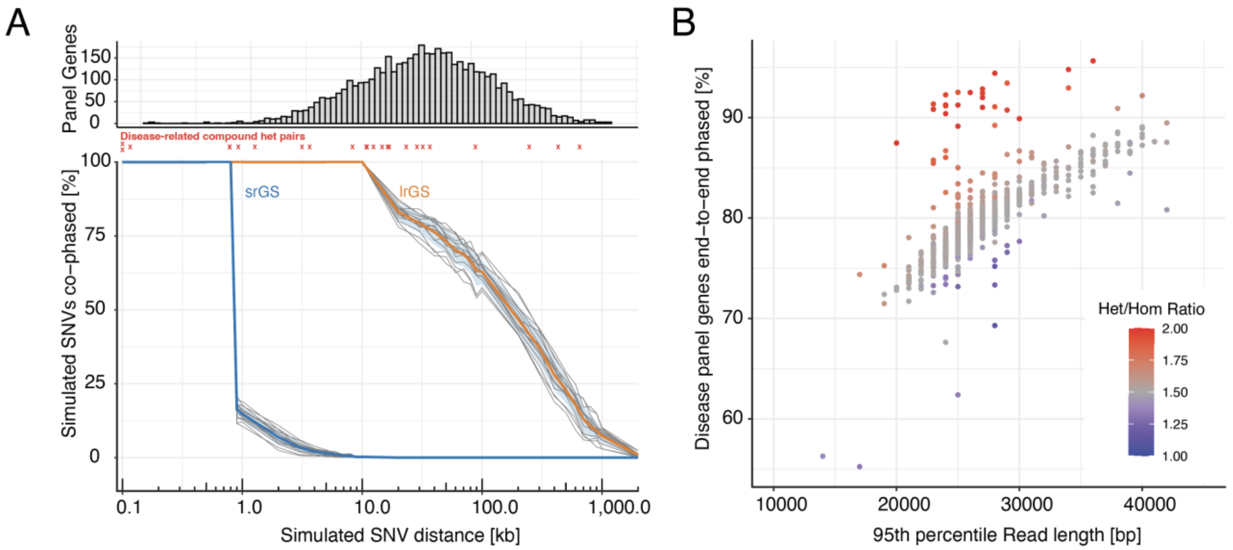

**A.** *Top*: Histogram of panel gene lengths (log scale). *Bottom*: Percentage of simulated SNV pairs that are phaseable (y-axis) as a function of inter-SNV distance. Results are shown separately for srGS (blue) and lrGS (orange). Red crosses mark distances of disease-related compound heterozygous variant pairs observed in our data. **B.** Scatter plot showing, per sample, the percentage of disease panel genes that are end-to-end phased (y-axis) versus the 95th percentile read length. Points are colored by the heterozygous-to-homozygous variant ratio (Het/Hom Ratio; higher ratio indicating more diverse ancestry).

**Figure S4: Visualization of 5mC data in IGV for four index cases tested for Prader-Willi and Angelman imprinting disorders.**

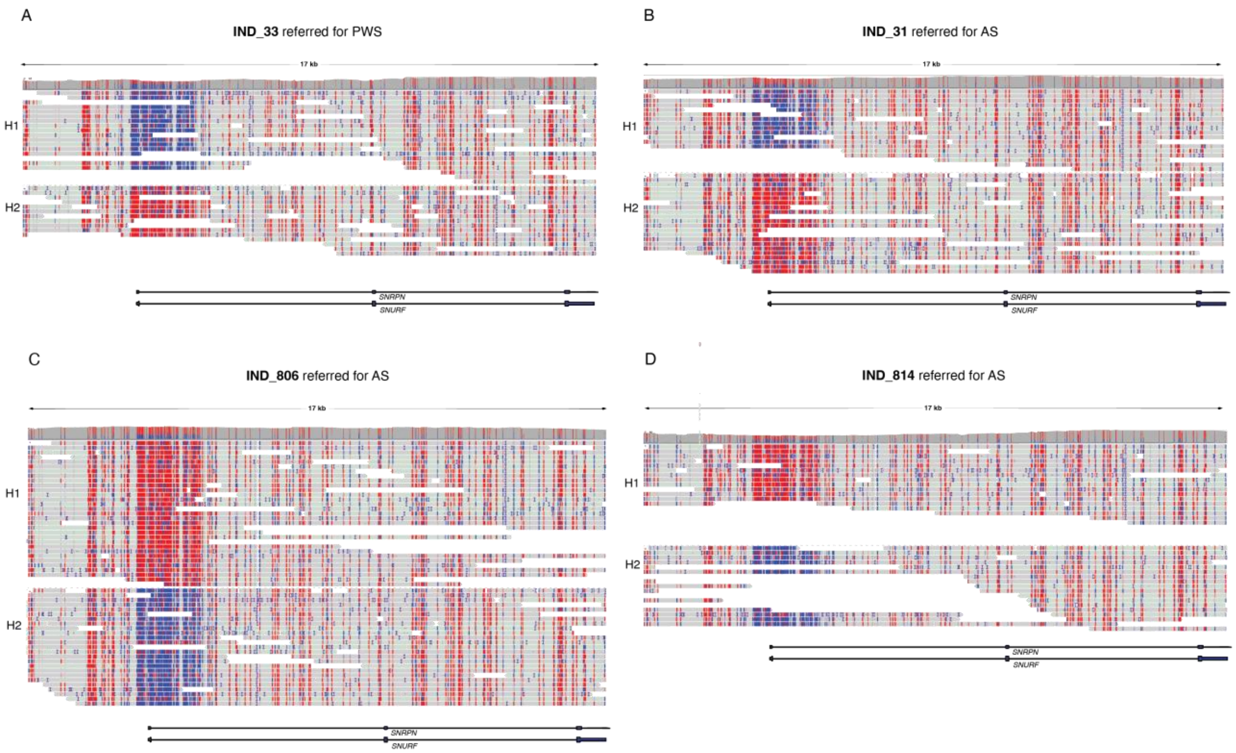

Four cases were referred for testing Prader-Willi Syndrome (PWS) or Angelman syndrome (AS) (panels **A-D**). Genetic confirmation of a clinical diagnosis can be obtained by methylation analysis of the *SNRPN/SNURF* locus on Chromosome 15. For all four cases, however, one hypermethylated haplotype (red) and one hypomethylated haplotype (blue) were detected, indicating the normal imprinting pattern, excluding the genetic diagnosis for PWS or AS. These results from lrGS are concordant for those obtained in SoC testing.

**Figure S5: Number of variants for interpretation after routine diagnostic filtering steps.**

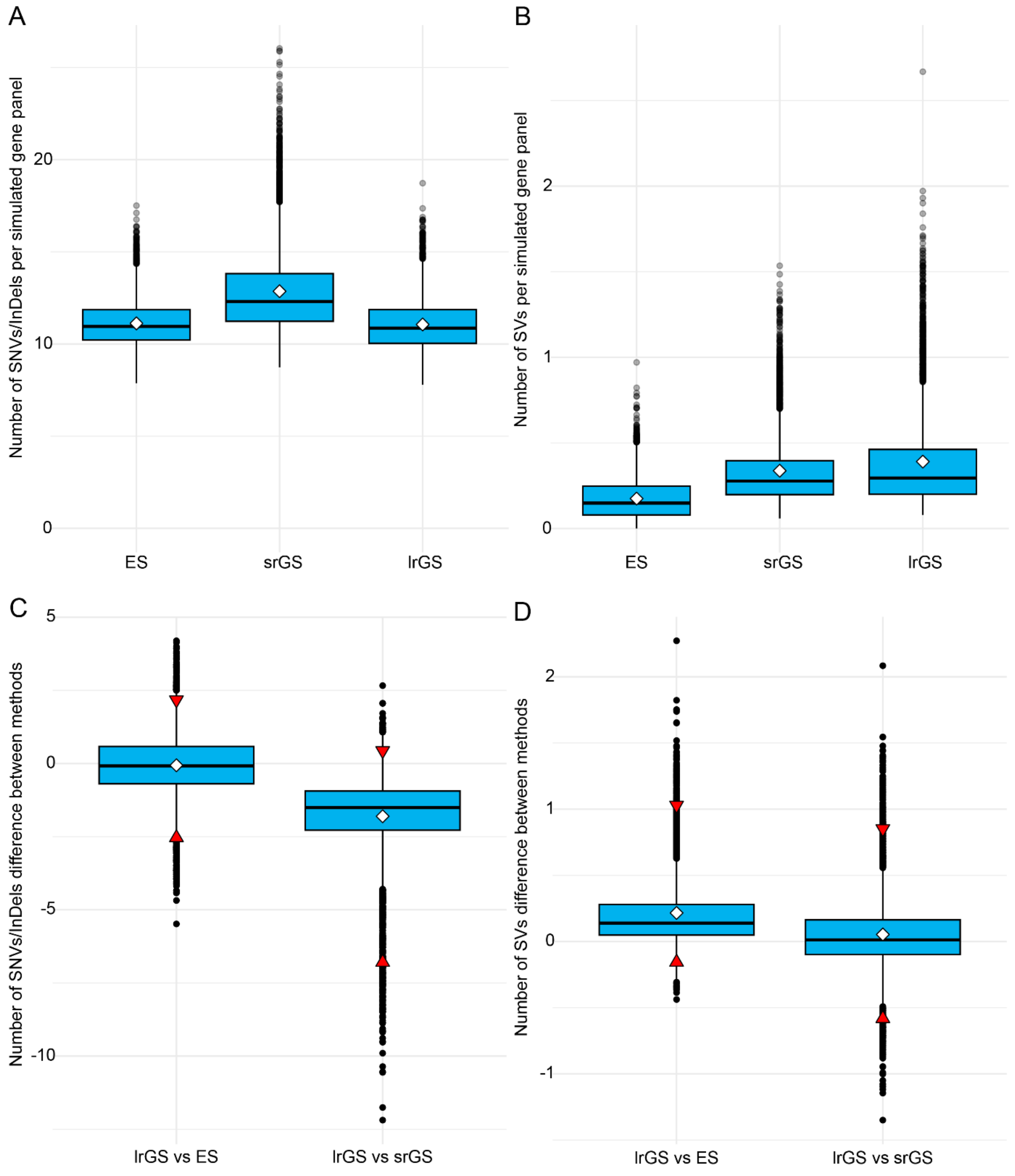

**A**) Mean number of SNVs and InDels identified by each method per simulated disease-gene panel of average size (n genes = 187). White diamonds indicate the mean **B**) Mean number of SVs identified by each method per simulated disease-gene panel. We additionally computed paired differences between the vectors of per-panel variant means of ES and lrGS, and of srGS and lrGS for SNV/InDels (**C**) and SVs separately (**D**). For each simulated panel, we calculated the difference between method 1 and method 2, generating a total of four difference vectors. The mean of each difference vector represents the typical per-panel difference in variant yield between methods. To characterize the expected variability across gene-panel compositions, we computed the 2.5th and 97.5th percentiles of the empirical distribution of method and variant type-specific differences. These percentiles define the 95% empirical range, which reflects the spread of differences attributable to panel composition. Red triangles pointing up represent the 2.5$^{th}$ percentile, and red triangles pointing down represent the 97.5$^{th}$ percentile. **C**) LrGS identified on average 0.06 fewer SNVs and InDels than ES, with 95% of the simulated panels ranging from 2.5 fewer to 2.2 more variants. In contrast, lrGS identified 1.8 fewer SNVs and InDels than srGS, and 95% of the panels ranged between 6.8 fewer and 0.4 more variants. **D**) LrGS identified 0.22 more SVs than ES on average, with 95% of the panels ranging from 0.16 fewer to 1.0 additional SV. Finally, lrGS identified 0.05 more SVs than srGS on average, with 95% of panels ranging from 0.58 fewer to 0.85 more SVs.

**Figure S6: Genes with impaired coverage**

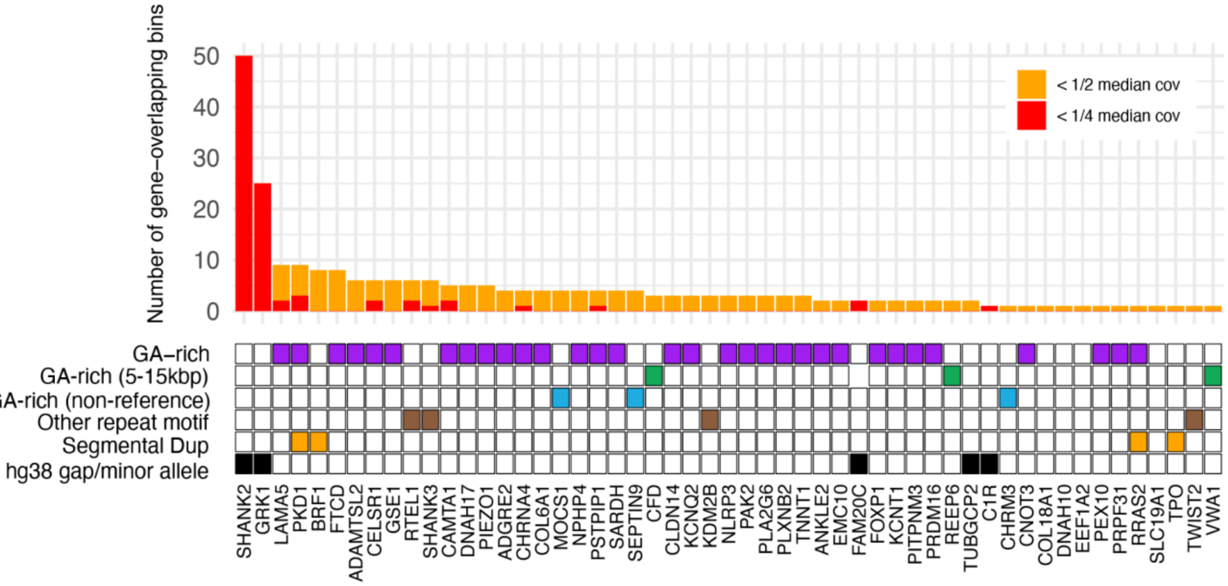

Bar plot showing genes with at least one low-coverage bin (<50% median coverage in >90% of samples; orange bars). Twelve genes contain bins with <25% coverage (red bars). The bar height indicates the fraction of each gene overlapping low-coverage bins. Squares below each bar denote whether at least i) one low-coverage bin overlaps an GA-rich region (purple), ii) is located between 5 and 15 kb from an GA-rich region (green), iii) carries GA-rich insertions not found on the reference (blue), iv) overlaps a repeat region with a non-GA motif (brown), v) a segmental duplication (orange) or vi) reference gap in GRCh38/hg38 (black).

**Figure S7: Examples of two GA-rich regions with <25% coverage that overlap medically relevant genes**

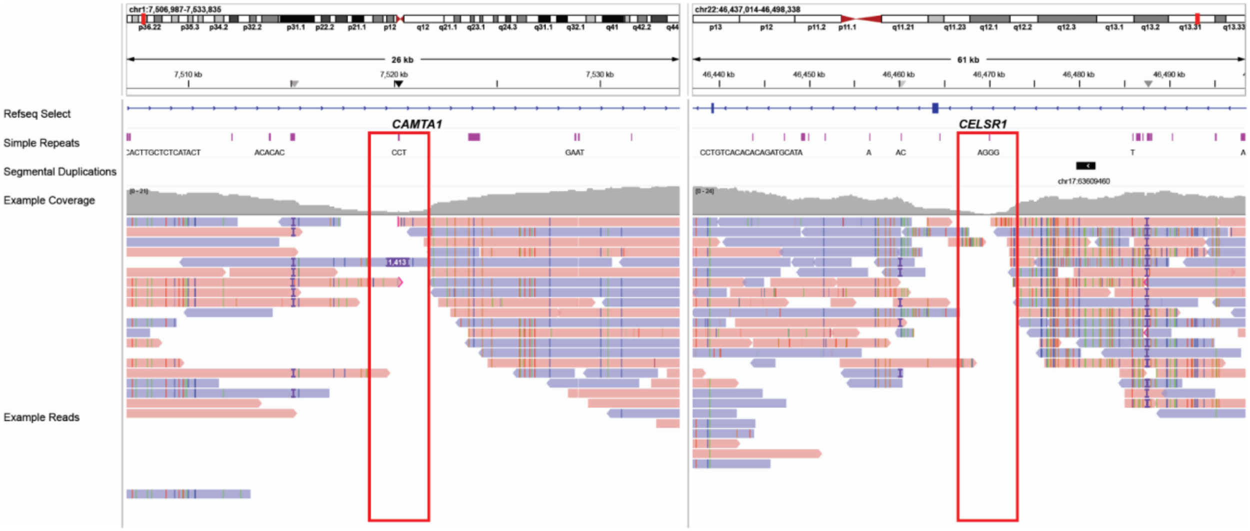

IGV screenshot of read mappings in a representative sample, showing two GA-rich regions with <25% coverage that overlap medically relevant genes. Red boxes indicate the GA-rich regions.

**Figure S8: Effect of GA-rich regions on read coverage**

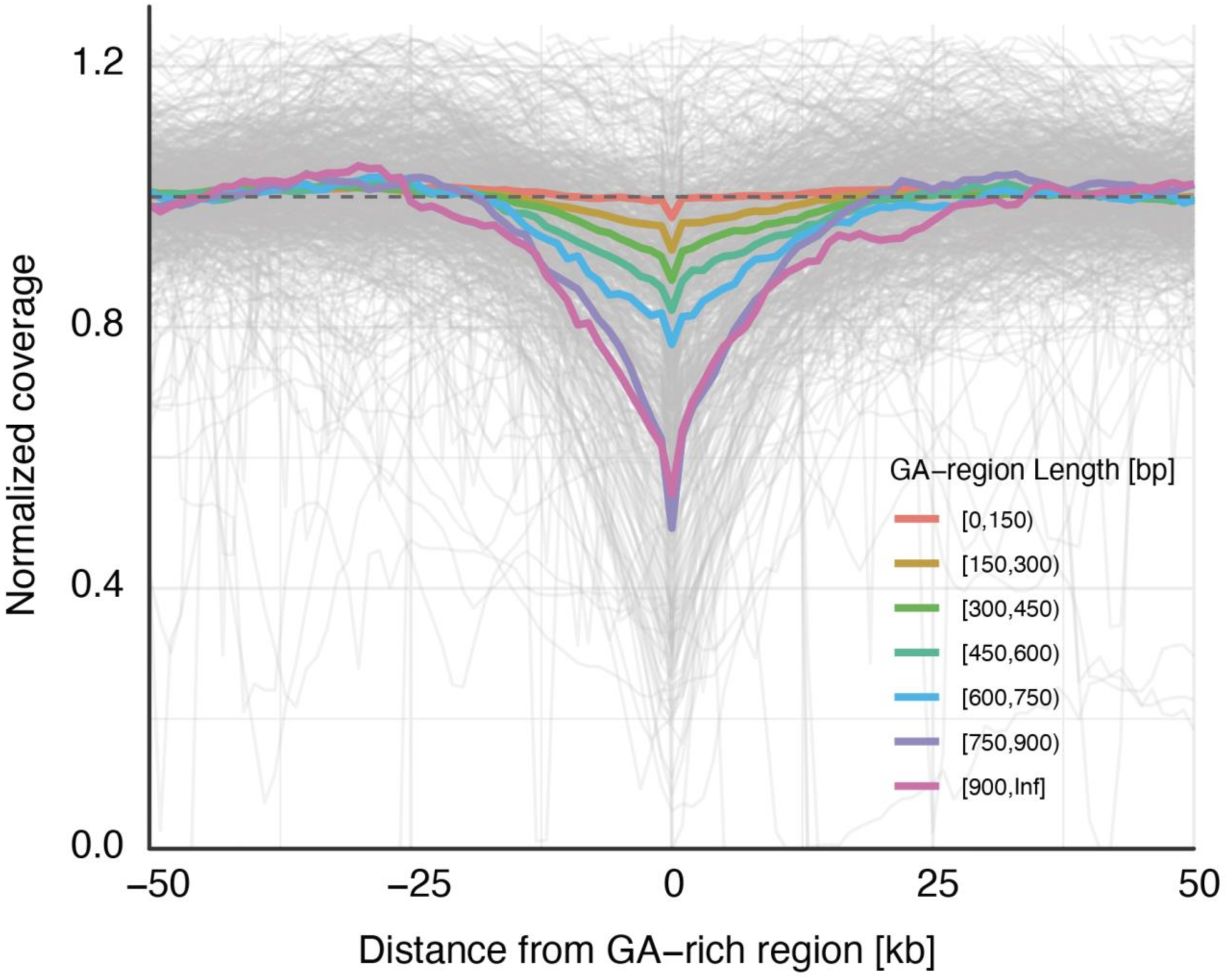

A stacked line plot illustrating the effect of GA-rich regions on read coverage within the GA-region and across a ±100 kb surrounding window. Coverage within the GA-region is shown at position 0 on the x-axis (‘Distance from GA-region’ = 0). Different colors indicate the median read coverage profiles for GA-regions of varying lengths. Each line represents the median coverage calculated across (1) 1,000 samples and (2) all GA-regions within the same size range. The strength of the dropout effect correlates with GA-region length.

**Figure S9: Coverage drops around individual GA-rich repeats**

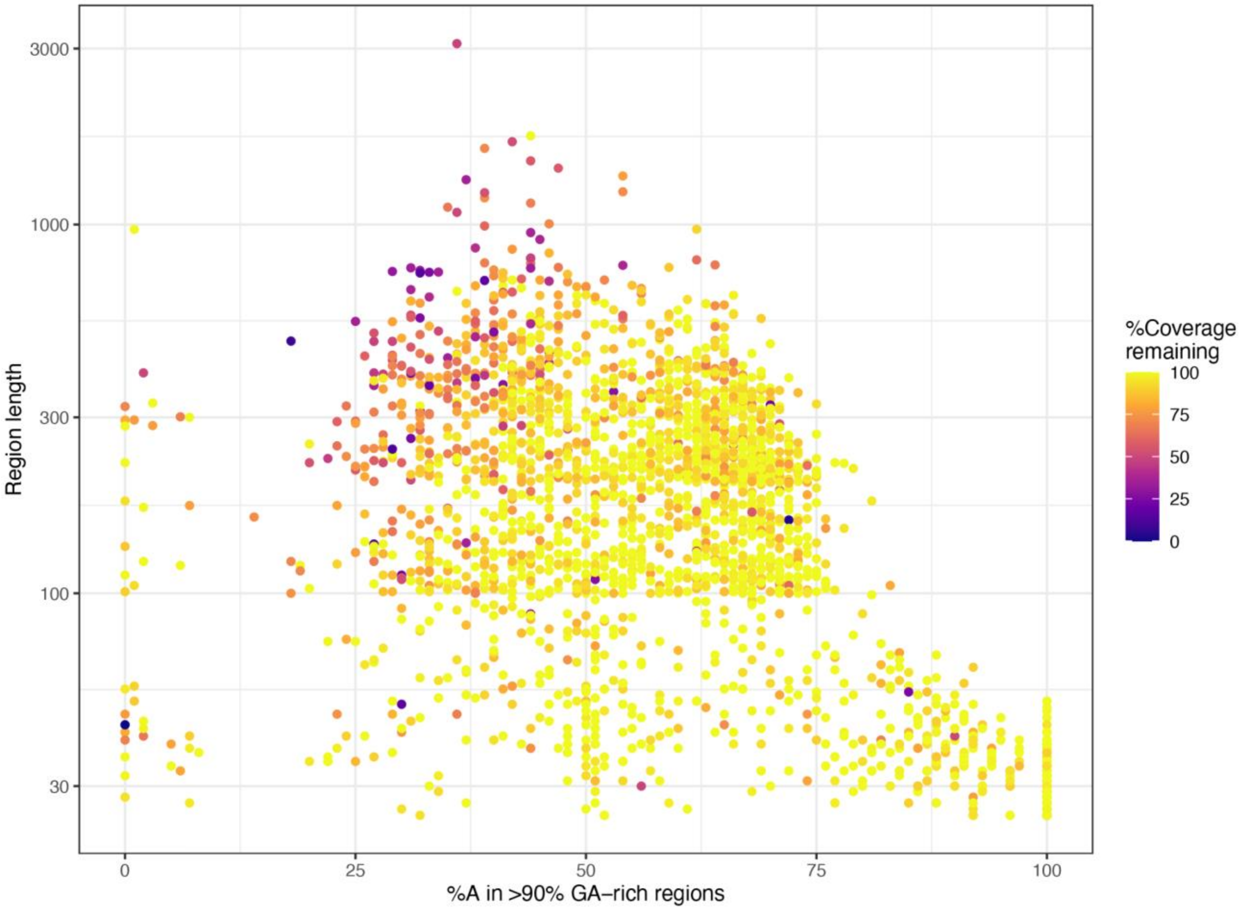

Scatter plot showing the strength of coverage drops around individual GA-rich repeats. Each point represents an annotated GA-rich region in hg38 longer than 30 bp. The x-axis shows %A content, and the y-axis shows GA-region length. Point color indicates the median coverage across 1,000 samples in the ±5 kb flanking regions (excluding the GA-rich region itself). The drop of coverage is strongest for regions containing <50% adenine.

**Figure S10: Titration analyses**

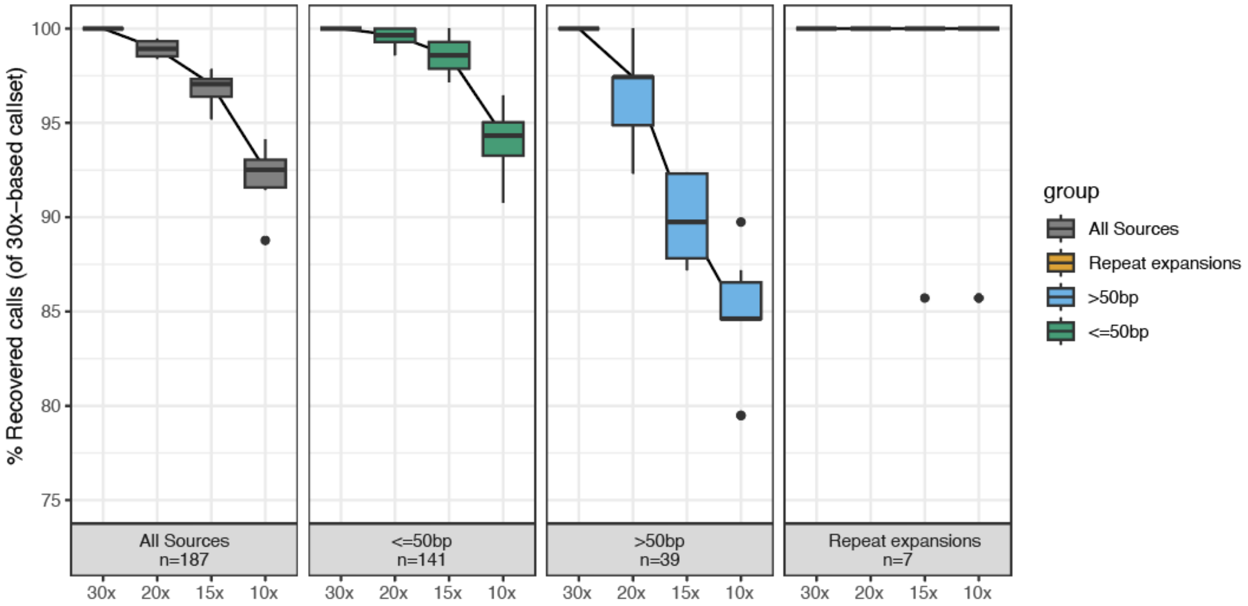

Results of re-calling all detected (likely) pathogenic variants on artificially downsampled genome-wide coverage levels (20x, 15x, 10x), stratified by variant type. The boxplots are generated from 10 random subsets of reads sampled from the initial 30x-coverage datasets.

**Figure S11: SoC referrals in 2024**

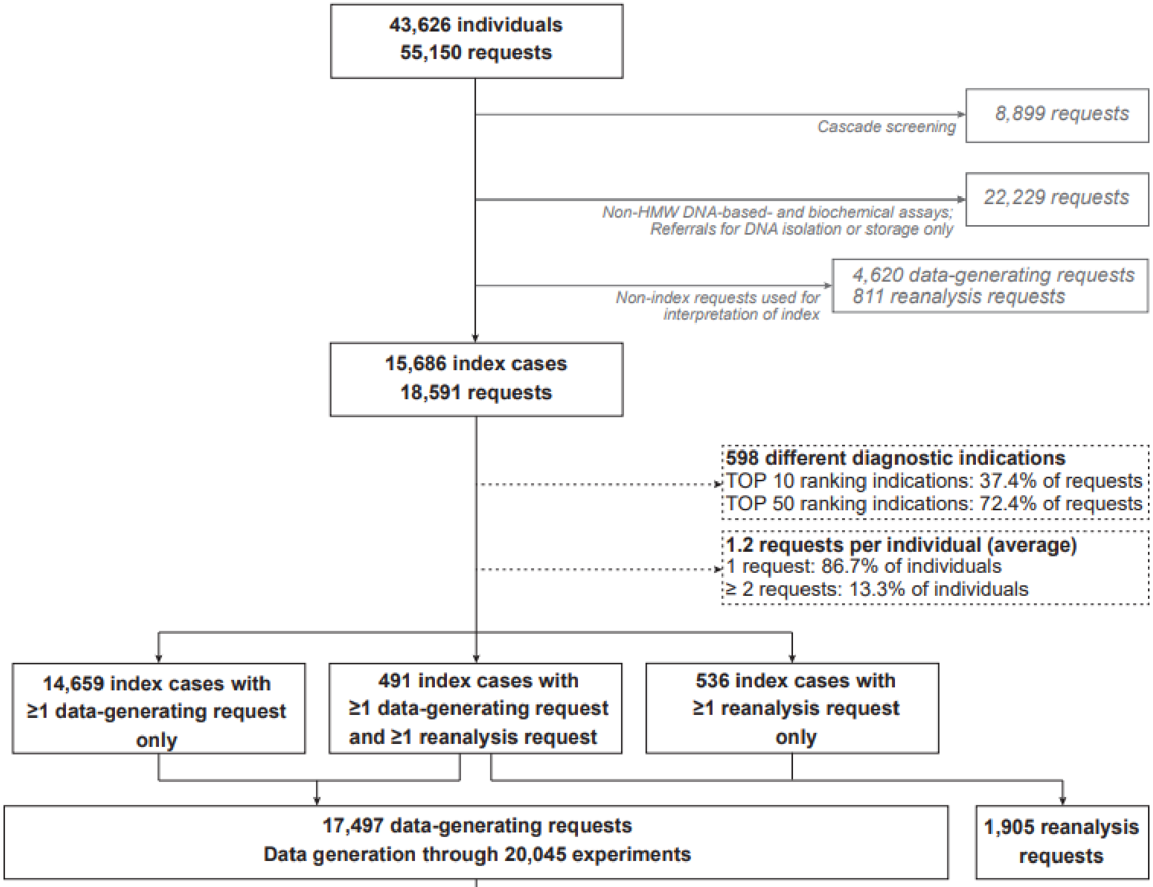

*Experiments per workflow (n)*

| Karyo-typing | FISH | srGS | ES | LR-amplicon | Array | QF-PCR | MLPA | Fragment | delPCR | tNGS | Sanger |
|---|---|---|---|---|---|---|---|---|---|---|---|
| 785 | 46 | 3,370 | 7,584 | 833 | 330 | 29 | 1,574 | 1,577 | 33 | 1,709 | 1,905 |

*Reported variants per workflow per variant type (n variants)*

| Variant type | Karyo-typing | FISH | srGS | ES | LR-amplicon | Array | QF-PCR | MLPA | Fragment | delPCR | tNGS | Sanger |
|---|---|---|---|---|---|---|---|---|---|---|---|---|
| Aneuploidy | 56 | 7 | 12 | 24 | 0 | 7 | 11 | 1 | 0 | 0 | 0 | 0 |
| SV | 37 | 10 | 8 | 5 | 1 | 0 | 0 | 0 | 0 | 1 | 0 | 0 |
| CNV | 11 | 6 | 191 | 235 | 5 | 38 | 0 | 51 | 0 | 0 | 8 | 0 |
| CN-LOH | 0 | 0 | 3 | 2 | 0 | 1 | 0 | 0 | 0 | 0 | 0 | 0 |
| SNV | 0 | 0 | 809 | 1,404 | 104 | 0 | 0 | 0 | 3 | 0 | 246 | 271 |
| InDel | 0 | 0 | 240 | 497 | 6 | 0 | 0 | 0 | 7 | 0 | 64 | 77 |
| Repeat expansion | 0 | 0 | 4 | 3 | 0 | 0 | 0 | 0 | 171 | 0 | 0 | 0 |
| UPD | 0 | 0 | 1 | 1 | 0 | 0 | 0 | 0 | 0 | 0 | 0 | 0 |
| Methylation | 0 | 0 | 0 | 0 | 0 | 0 | 0 | 0 | 2 | 0 | 0 | 0 |
| mtDNA (SNV +CNV) | 0 | 0 | 14 | 4 | 69 | 0 | 0 | 0 | 0 | 0 | 0 | 0 |
| *Total* | **104** | **23** | **1,282** | **2,175** | **185** | **46** | **11** | **52** | **183** | **1** | **318** | **348** |

**Figure S12: Detailed insights in modeled transitions in diagnostic outcome as a consequence of a generic lrGS-first approach.**

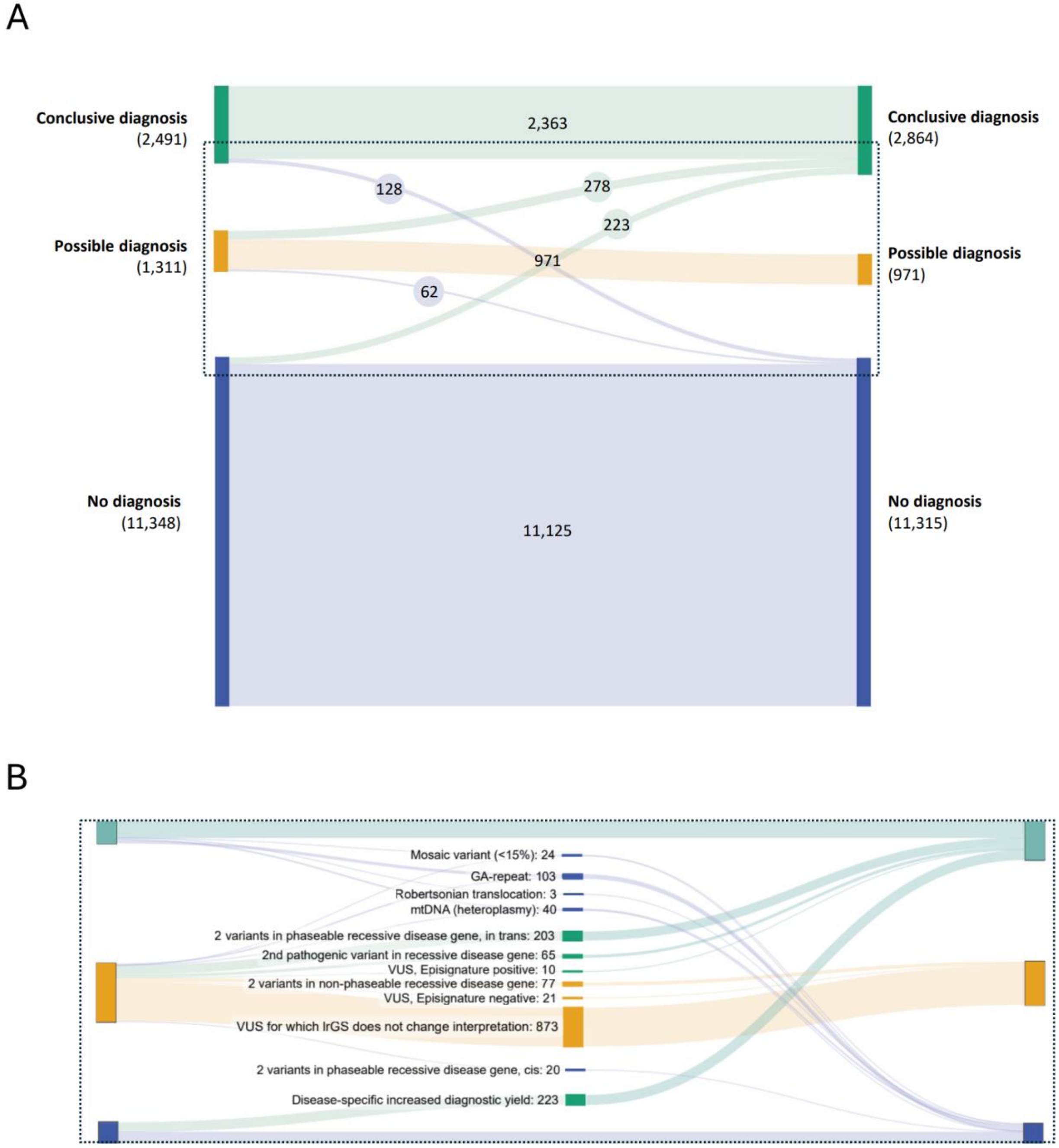

For each index case in 2024, we modeled the impact of generic lrGS-first approach. **A**) Cohort level overview of shifts in diagnostic outcome. The dotted box is represented in more detail in (B) to highlight the modeled changes in outcome. **B**) For 103 index cases (n=66 conclusive diagnoses and n=37 possible diagnoses), a GA-based repeat was reported, which would not be identified in a lrGS-first approach. Three index cases were reported to have an SV that was mediated by a repeat that exceeds the HiFi read length (n=3 conclusive diagnoses by Robertsonian translocations). Lastly, we investigated the reports mentioning mtDNA variants (any level of heteroplasmy, n=40, including 39 conclusive, and 1 possible diagnosis, respectively) and germ-line derived mosaics (<15%, n=24, including 20 conclusive and 4 possible diagnoses, respectively), and predicted that none of these would be detected. We next modeled the positive impact of a lrGS-first approach on diagnostic yield. For this, we used our observations from our parallel study, as well as from large scale publications reporting on the added diagnostic value of lrGS in specific disease and leveraged this information to the 1,311 possible diagnoses. First, we incorporated the use of variant phasing in recessive disease genes, while considering the phase-ability of the respective genes. For 300 index cases, possible diagnoses were reported in recessive disease genes, which would require segregation analysis of family members in SoC. For 223 of them, variants were in genes for which phasing would be directly possible from lrGS. From our model, 91.3% (n=203) would be in *trans* (e.g. conclusive diagnosis) and 8.7% (n=20) in *cis* (e.g. no diagnosis). For the remaining 77 cases, variants were in genes challenging to phase start-to-end, maintaining their possible diagnosis status. Second, we modeled the advantage of lrGS to uncover a (likely) pathogenic variant in an index for whom a single mono-allelic variant was already identified. In this cohort, for 152 index cases, such a mono-allelic (likely) pathogenic allele was reported. Extrapolating from our own observations and previous studies[14,15], the chance to uncover such a 'missing second allele' is 42.9%, suggesting that for 65 of these indexes a conclusive is to be expected. Third, we focused on (dominant) variants of unknown significance (VUS) in genes or loci for which pathogenicity can be established by testing aberrant DNAm profiles. In total, 31 index cases were reported to have such VUS and for whom such read-out would be beneficial. Prior studies show that 32.4% of VUS can be reclassified to pathogenic variants, which translates to 10 conclusive diagnoses, and 21 that remain a possible diagnosis. Lastly, we predicted the increased diagnostic yield from the discovery of so far hidden variation in index cases in whom the genetic diagnoses remained unresolved. For this, we matched the reported disease-specific increase from large scale lrGS studies to our diagnostic requests. This resulted in a predicted shift of 223 index cases from no diagnosis to conclusive diagnosis.

**Figure S13: Illustration of the geometric framework for estimating the probability that two heterozygous SNVs can be co-phased.**

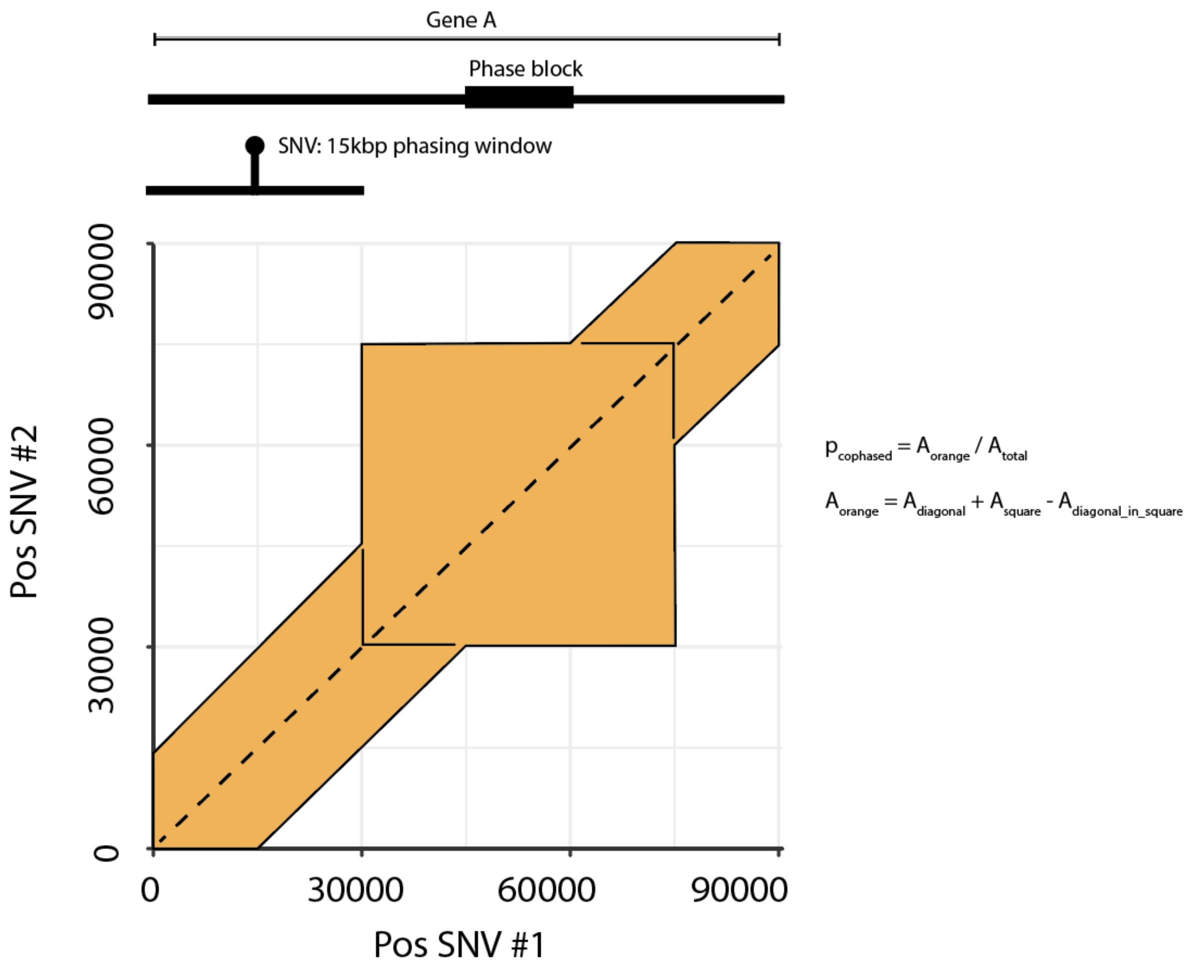

Illustrated is the example case of a 90 kb gene with one pre-existing 15 kb phase block from position 45–60 kb. Any SNV pair can be represented as a point on a two-dimensional plane, where the x- and y-coordinates denote their positions along the gene. A pair of SNVs can be co-phased in two ways: (a) if the two SNVs lie sufficiently close to be spanned by a single read, or (b) if both fall within or in read-length range to the same pre-existing phase block. In this geometric analogy, case (a) corresponds to the diagonal band around x=y with thickness 2*(read length), and case (b) corresponds to one or more squares centered around the diagonal. The probability that two heterozygous SNVs can be co-phased is given by the fraction of the total plane area covered by these regions (shaded orange). The areas of both regions are functions of the read length, phase-blocks, and gene length. The illustration shows a simple case with one phase block. The calculation can be performed analogously for any number and configuration of phase blocks.

#### Tables

Tables are provided in Supplementary Appendix 2 (.xlsx spreadsheet)

**Table S1: Sequencing cohort of 1000 samples including SoC tests and outcome**

**Table S2: Sequencing characteristics and number of variants per variant type**

**Table S3: Variant details**

**Table S4: Phasing information per disease gene**

**Table S5: Results of skewed X-inactivation analysis**

**Table S6: Results of DNAm profile analysis**

**Table S7: Interpretation burden**

**Table S8: Genes with impaired coverage**

**Table S9: Titration analysis**

**Table S10: Model assumptions used in the impact analysis**

**Table S11: Overview of literature used in the impact analysis**

**Table S12: Reported variants per workflow in SoC (2024 diagnostic referrals) and recall rate in lrGS used in the impact analysis**

**Table S13: Prioritization and interpretation criteria for the different variant types**

**Table S14: Disease genes**

15. AlAbdi L, Shamseldin HE, Khouj E, et al. Beyond the exome: utility of long-read whole

genome sequencing in exome-negative autosomal recessive diseases. Genome Med 2023;15(1):114.
